# Two year data from a preliminary study of double lesion site MRgFUS treatment of Essential Tremor targeting the thalamus and the posterior subthalamic area

**DOI:** 10.1101/2020.12.27.20248723

**Authors:** Ayesha Jameel, Wladyslaw Gedroyc, Dipankar Nandi, Bryn Jones, Olga Kirmi, Sophie Molloy, Yen Tai, Gavin Charlesworth, Peter Bain

**Affiliations:** Department of Radiology, Imperial College Healthcare NHS Trust, London, UK; Department of Neurosciences, Imperial College Healthcare NHS Trust, London, UK; Department of Neurosciences, Division of Brain Sciences, Imperial College London, UK

**Keywords:** MR-guided focused ultrasound, Essential tremor, Zona Incerta, Ventralis Intermedius nucleus, Bain-Findley Spiral

## Abstract

**Background:** MR-guided focused ultrasound (MRgFUS) is an effective treatment for essential tremor (ET). However, the optimal intracranial target sites remain to be determined.

**Objective:** To assess MRgFUS induced sequential lesions in (anterior-VIM/VOP nuclei) the thalamus and then posterior subthalamic area (PSA) performed during the same procedure for alleviating ET.

**Methods:** 14 patients had unilateral MRgFUS lesions placed in anterior-VIM/VOP then PSA. Bain-Findley Spirals were collected during MRgFUS from the treated arm (BFS-TA) and throughout the study from the treated (BFS-TA) and non-treated (BFS-NTA) arms and scored by blinded assessors. Although, the primary outcome was change in the BFS-TA from baseline to 12 months we have highlighted the 24 month data.

Secondary outcomes included the Clinical Rating Scale for Tremor (CRST), Quality of Life for ET (QUEST) and PHQ-9 depression scores.

**Results:** The mean improvement in the BFS-TA from baseline to 24 months was 41.1% (p<0.001) whilst BFS-NTA worsened by 8.8% (p<0.001). Intra-operative BFS scores from the targeted arm showed a mean 27.9% (p<0.001) decrease after anterior-VIM/VOP ablation and an additional 30.1% (p<0.001) reduction from post anterior-VIM/VOP to post-PSA ablation.

Mean improvements at 24 month follow-up in the CRST-parts A, B and C were 60.7%, 30.4% and 65.6% respectively and 37.8% in QUEST-tremor score (all p<0.05). Unilateral tremor severity scores decreased in the treated arm (UETTS-TA) 72.9% (p=0.001) and non-treated arm (UETTS-NTA) 30.5% (p=0.003). At 24 months residual adverse effects were slight unsteadiness (n=1) and mild hemi-chorea (n=1).

**Conclusion:** Unilateral anterior-VIM/VOP and PSA MRgFUS significantly diminished contralateral arm tremor with improvements in arm function, tremor related disability and quality of life, with an acceptable adverse event profile.

## Introduction

Essential Tremor (ET) is defined as an isolated tremor syndrome of bilateral upper limb action tremor, of at least 3 years duration, in which there can be tremor present in other parts of the body but other neurological signs, such as Parkinsonism, dystonia and ataxia are absent.^1^ The prevalence of ET is estimated to be approximately 2%, increasing to 4%-6% in people aged ≥ 40 years old.^2, 3^

It is estimated that 25%-55% of patients with ET are medication refractory.^4^ ET can produce substantial impairment of manual function and thus activities of daily living, resulting in disability and social handicap. Patients with ET are at increased risk of anxiety, which exacerbates tremor, and depression.^5, 6, 7, 8^

An evidence based review concluded that for appropriate patients with medically refractory ET, or intolerant of anti-tremor medications, treatment with unilateral Ventralis Intermedius (VIM) thalamic DBS or radiofrequency or MRgFUS thalmotomy was possibly helpful. The authors of the review considered there to be insufficient evidence to recommend bilateral VIM DBS or unilateral gamma-knife therapy. ^9^

MR guided focused ultrasound (MRgFUS) is a non-invasive treatment for ET. By utilising high intensity focused ultrasound waves, generated from a multi-element transducer and focused onto a small target, accurate thermal ablation of the target brain tissue can be achieved under real time image guidance. MRgFUS utilises both MR-imaging for targeting and MR-thermal imaging to plan, monitor and ablate the target tissue under operator control whilst the patient is awake.

Currently, most centres treating ET with MRgFUS place lesions in the VIM nucleus of the thalamus (Hassler classification) ^10^ which receives the vestibular and proprioceptive muscle spindle afferent inputs. ^11,12,13,14,15^ Typically the lesion is sited 25% along the AC-PC line (i.e. 6-9mm anterior to PC depending on the AC-PC length), 0-2mm superior to the AC-PC plane and 8-12mm lateral to the lateral wall of the 3^rd^ ventricle.

Boutet et al. (2018), using this approach, elegantly demonstrated that the most effective lesions for ET suppression are placed in the posterior part of VIM close to its posterior boundary with the Ventralis Caudalis (VC) nucleus but that slight posterior encroachment on VC was associated with a 38 times greater incidence of contralateral sensory disturbance.^16^ This finding likely explains why a meta-analysis of MRgFUS treatment of ET showed that 15.3% of patients had persisting paraesthesia at 12 month follow up. ^17^

A randomised controlled trial of MRgFUS lesions in VIM versus sham MRgFUS demonstrated significant reduction in contralateral arm tremor over a 12 month period, with a resultant reduction of tremor related disability and improvement in quality of life for the patients receiving actual VIM lesions.^14^ Sham MRgFUS produced only a 1.25% reduction of tremor 3 months post-MRgFUS (i.e. a negligibable placebo effect).^14^

Stereotactic lesions (including radiofrequency thermal ablations and more recently MRgFUS lesions) in the cerebellothalamic tract (CTT), in the posterior subthalamic area (PSA) (which includes the zona incerta (ZI), posterior subthalamic nucleus and fibre connection area) have been demonstrated to alleviate tremor in the contralateral upper limb.^18,19^ This follows a trend for targeting this area in patients with ET using radiofrequency lesions and latterly DBS.^18^ There have been no studies comparing VIM versus other targets for treating people with ET using either radiofrequency lesions, DBS or MRgFUS.

Targeting anterior-VIM/VOP and posterior subthalamic area (PSA) in the same procedure has been the standard approach of our and the Oxford functional neurosurgery centres for over 2 decades of radiofrequency lesion and DBS surgery for ET. Both centres target anterior-VIM/VOP approximately 2.7mm-3.7mm anterior to the standard VIM target (25% of the AC-PC line, anterior to PC) depending on the length of the patient’s AC-PC line (24 to 28mm), with similar lateral and vertical co-ordinates, so that the lesion is likely to straddle Ventralis Oralis Posterior (VOP) and the anterior part of VIM.^(20,21)^ This paper describes our experience of unilateral MRgFUS in patient with medically refractory ET, sequentially treating anterior-VIM/VOP and then PSA in the same procedure.

## Methods

The study was approved by the Research Ethics Committee (REC no: 15LO1538). The unilateral MRgFUS procedures were performed between July 2016 and November 2017. In each case, during a single procedure, the anterior-VIM/VOP was the area targeted first and then, because of varying degrees of residual tremor, the target was moved manually (based on the neuroimaging) and the PSA was lesioned. In each case the clinical decision to lesion PSA was pragmatic and based on the views of the patient and the neurologist (PGB) that the residual tremor post the anterior-VIM/VOP thalamotomy was still intrusive or could be improved. We also considered whether a lesion in PSA would add longevity to the benefit caused by the anterior-VIM/VOP lesion.

### Patient Selection

Patients aged more than 21 years with moderate or severe ET causing significant disability and with an inadequate response to 2 or more anti-tremor medications were eligible for the study providing they met the inclusion criteria (Appendix 1) and had no parameters on the exclusion criteria (Appendix 1). Minimum tremor severity for trial entry was a postural or intention tremor score of ≥ 2 on the Clinical Rating Scale for Tremor (CRST) (Appendix 2) in the dominant arm, with a minimum tremor related disability cut off score of ≥ 2 on the disability subsection (Part C) of the CRST. ^22^

### MRgFUS treatment

The procedural methodology is described in detail in Appendix 3. Key points regarding our approach to initial targeting of the anterior-VIM/VOP and PSA are given below and in Figure 1a.

**Figure 1:**
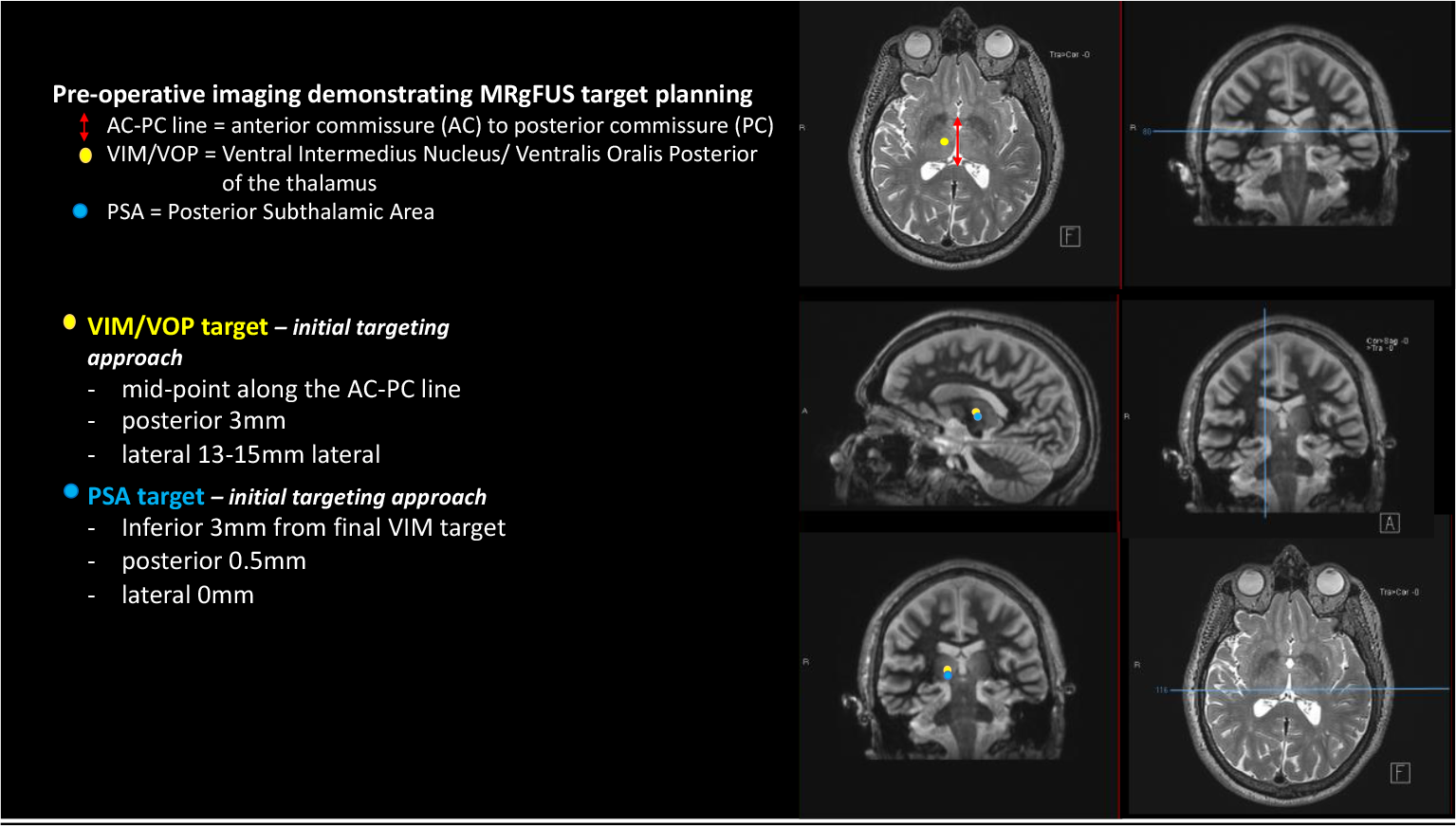
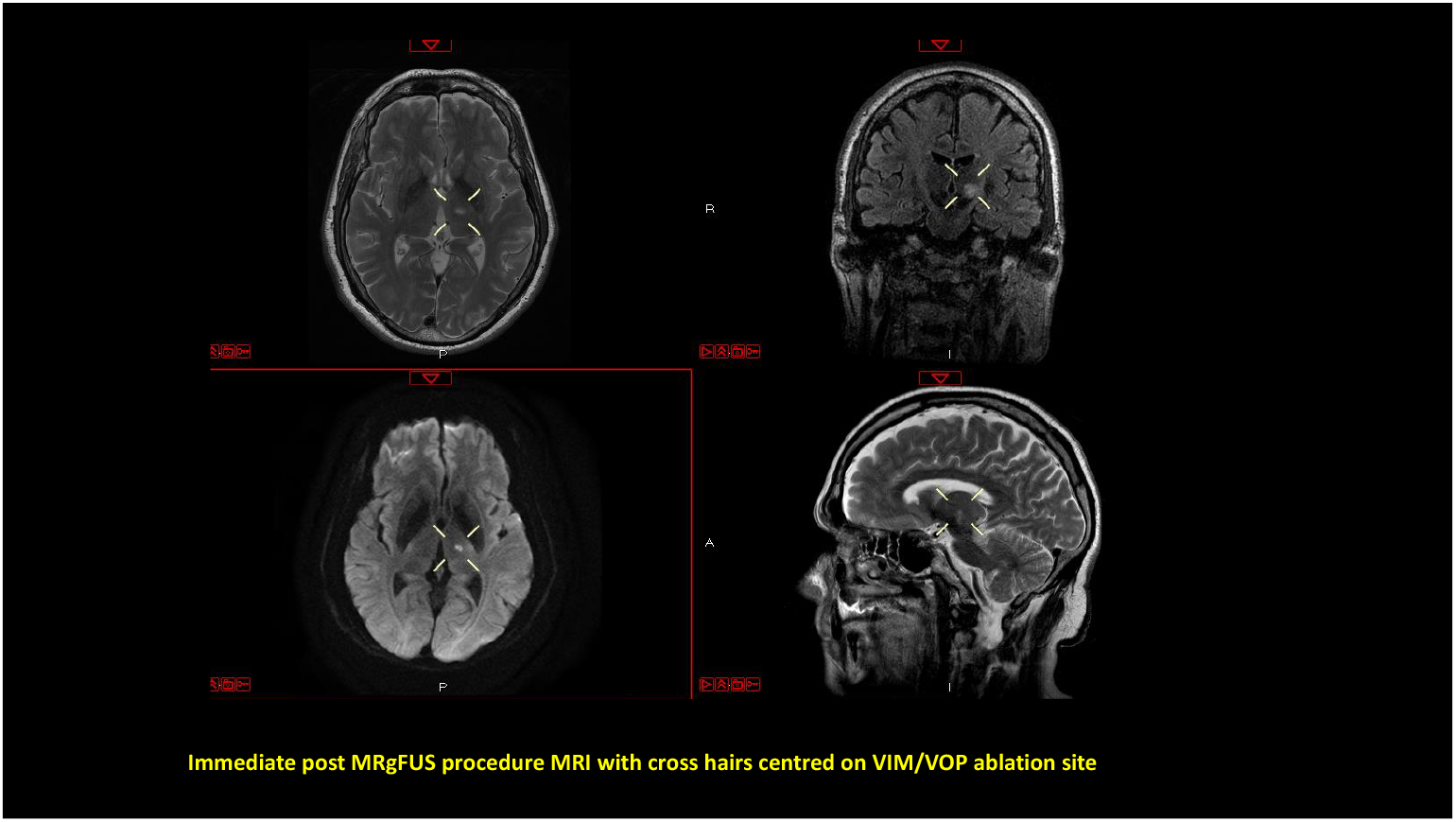
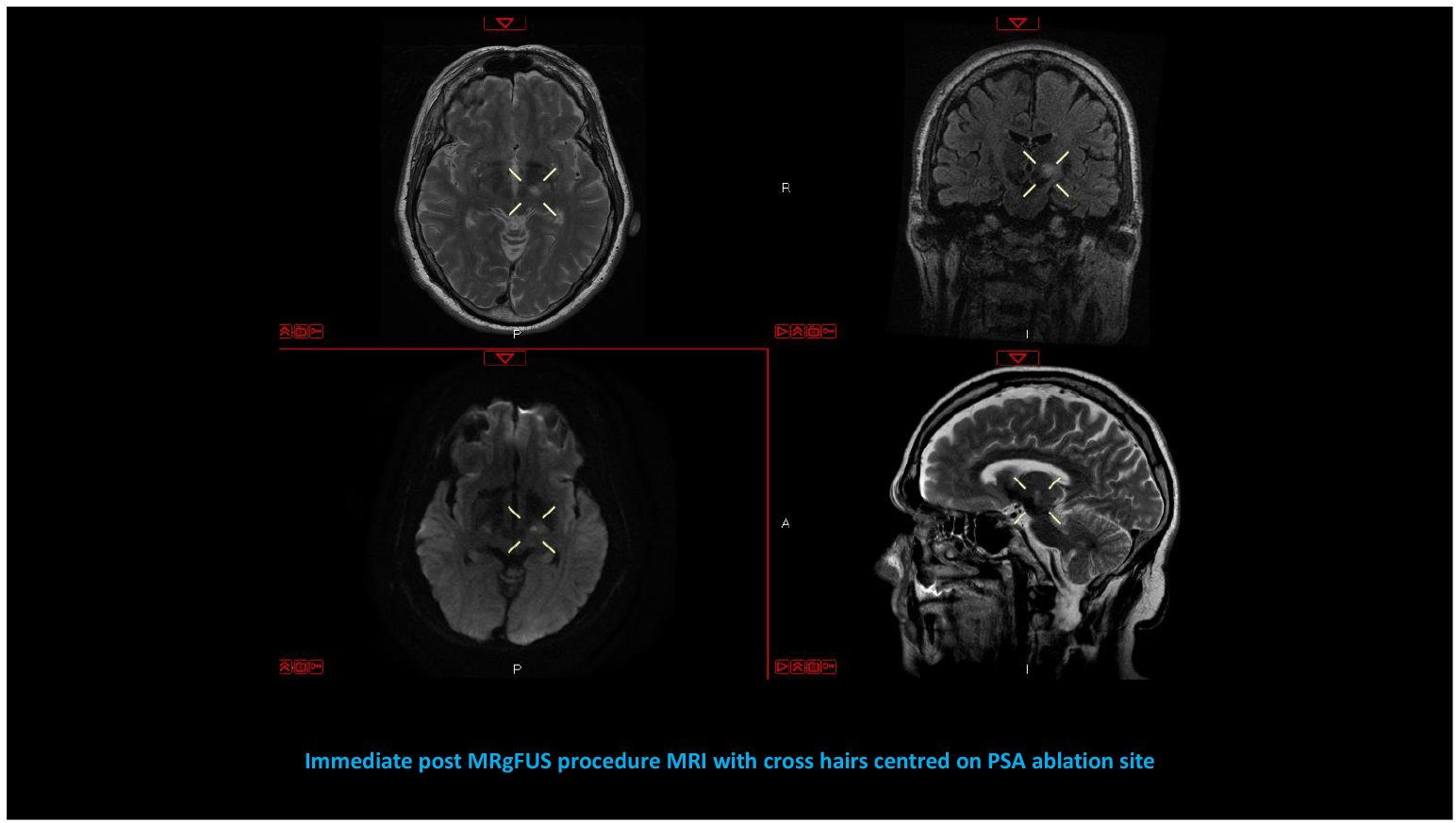
Targeting the VIM/VOP and ZI. ➢ 1a: Annotated pre-procedure MRI demonstrating approach to the Thalamic and PSA targets ➢ 1b-1: Post MRgFUS procedure MRI demonstrating VIM/VOP ablation ➢ 1b-2: Post MRgFUS procedure MRI demonstrating PSA ablation

### Identifying the Thalamic and PSA targets

Using the preoperative MRI study, stereotactic planning of the target was performed. The anterior-VIM/VOP was initially targeted at 3mm posterior and 13-15mm lateral to the mid-point along the AC-PC line (Figure 1a). This target was then cross correlated with the distance from the third ventricle and the internal capsule. The optimal distance from the wall of the third ventricle is 11-11.5mm and from the lateral border of the thalamus is 2mm.

Trial and treatment sonications (see Appendix 3) were performed to ensure tremor suppression was achieved by tailoring targeting to individual neuroanatomy and sonication parameters to individual neuromodulatory response, as such the degree of movement from the initial target to the final target for each nucleus is patient specific. Once a satisfactory lesion was placed in anterior-VIM/VOP then, providing a clinically significant tremor remained in the treated hand, the target was switched to the PSA. The rationale for using the PSA is supported by published literature from DBS treatment of ET.^40^ The location of the PSA varies slightly between patients (as is true of all the common functional stereotactic targets in clinical practice). Our standard approach to PSA targeting was movement 3 mm inferior, 0.5mm posterior and 0.0mm lateral to the anterior-VIM/VOP target (Figure 1a).

### Patient Assessments

Following screening eligible patients were assessed at baseline (within 14 days of MRgFUS), and then at 1 day, 1 week, 1, 3, 6, 12 and 24 months post-procedure.

### Outcome measures

#### Primary outcome

The primary outcome measure was the Bain & Findley Spiral (BFS) score in the treated arm at 12 months post-MRgFUS compared to baseline. As the study was extended to 24 months we have reported both 12 & 24 months data and emphasized the latter as it is of greater importance. The BFS is a reliable and validated method of assessing ET in which spirals are scored from 0 (no tremor) to 10 (very severe).^(23, 24)^

Freehand spirals were collected from both hands of patients at screening, baseline and the 1, 3, 6, 12, and 24 month visits. The intra-operative spirals were serially collected from the target hand prior to the first and then after each sonication. The patients drew on a board held above them whilst lying supine on the scanner. These two collections of spirals were separated, anonymized, and presented in random order to three blinded movement disorder neurologists (GC, YT, SM) to score separately using the BFS method.^23^

#### Secondary Outcome measures

The Clinical Rating Scale for Tremor (CRST) has been widely used in the field of MRgFUS to score tremor severity.^11, 13, 14, and 15.^ All components of the CRST were assessed unblinded by a tremor expert (PB).

The CRST is divided into 3 parts (Appendix 2):

- Part A scores whole body tremor severity considering anatomical location (arm, leg, voice etc.) and its components (rest, postural and intention tremor).
- Part B assesses tremor impact on specific motor tasks (e.g. drawing and pouring water).
- Part C assesses tremor related disability.

The effect of tremor on quality of life was documented using the Quality of life in Essential Tremor Questionnaire (QUEST) and depression monitored using the PHQ-9 questionnaire (Appendix 4 & 5 respectively). ^25, 26^

Within CRST Part A, the upper extremity total tremor score (UETTS) and its components: rest tremor (RT), postural tremor (PT) and intention tremor (IT) were also analysed for the treated and non-treated arms separately

#### Statistical methods

Statistical analysis was performed with SPSS version 25 using the non-parametric Wilcoxon Signed-Rank Sum test for paired data and the Friedman test for multiple data sets.

## Results

17 patients were screened, 2 failed entry criteria and 15 patients were enrolled in the study. One patient declined treatment and thus 14 patients underwent MRgFUS. The mean SDR was 0.43 (range: 0.35-0.51). In all 14 patients, the maximum temperatures achieved were within the desired thermal ablation range and there were successful lesion placements in both anterior-VIM/VOP and PSA (Figure 1b). One patient withdrew from the study after completing the one day post-MRgFUS assessment, having had an excellent response without adverse effects. The remaining 13 patients (8 male, 5 female) had a mean age of 69 years (range: 52-85 years), with a mean age at tremor onset of 32 years (range: 4-68) and mean duration of tremor of 38 years (range: 7-67). 11 had familial and 2 sporadic ET. 8 had head tremor and 7 vocal tremor in addition to bilateral upper limb tremor. The flow diagram for the study is shown in Appendix 6 and baseline characteristics of the patients’ tremors in Table 1:

**Table 1:** Baseline characteristics of patients’ tremor:

| **Baseline Tremor Scores** | **Mean (range)** |
| --- | --- |
| Bain Findley Spiral -Treated Arm (BFS-TA) | 5.3 (4-8) |
| Bain Findley Spiral - Non-Treated Arm (BFS-NTA) | 4.7 (3-7) |
| Clinical Rating Scale for Tremor Total (CRST) | 58 (35-83) |
| CRST Part A (CRST-A) | 16.5 (8-28) |
| CRST Part B (CRST-B) | 21.7 (12-30) |
| CRST Part C (CRST- C) | 19.8 (14-28) |
| Upper Extremity Total Tremor Score - Treated Arm (UETTS-TA) | 6.7 (6-9) |
| Rest Tremor - Treated Arm (RT-TA) | 0.7 (0-2) |
| Postural Tremor -Treated Arm (PT-TA) | 3.3 (2-4) |
| Intention Tremor - IT (IT-TA) | 2.9 (2-4) |
| Upper Extremity Total Tremor Score - Non-treated Arm (UETTS-NTA) | 5.8 (2-9) |
| Quality of Life in Essential Tremor – QUEST | 87.1 (56–108) |

### Primary Outcome

The change in the BFS in the treated arm (BFS-TA) was from 6.24 at baseline to 3.5 at 12 months, with a score of 3.7 at 24 months post MRgFUS (improvements of 43.5% and 41.1% from baseline respectively (both p<0.001, Friedman Test), (Figure 2a).

**Figure 2:**
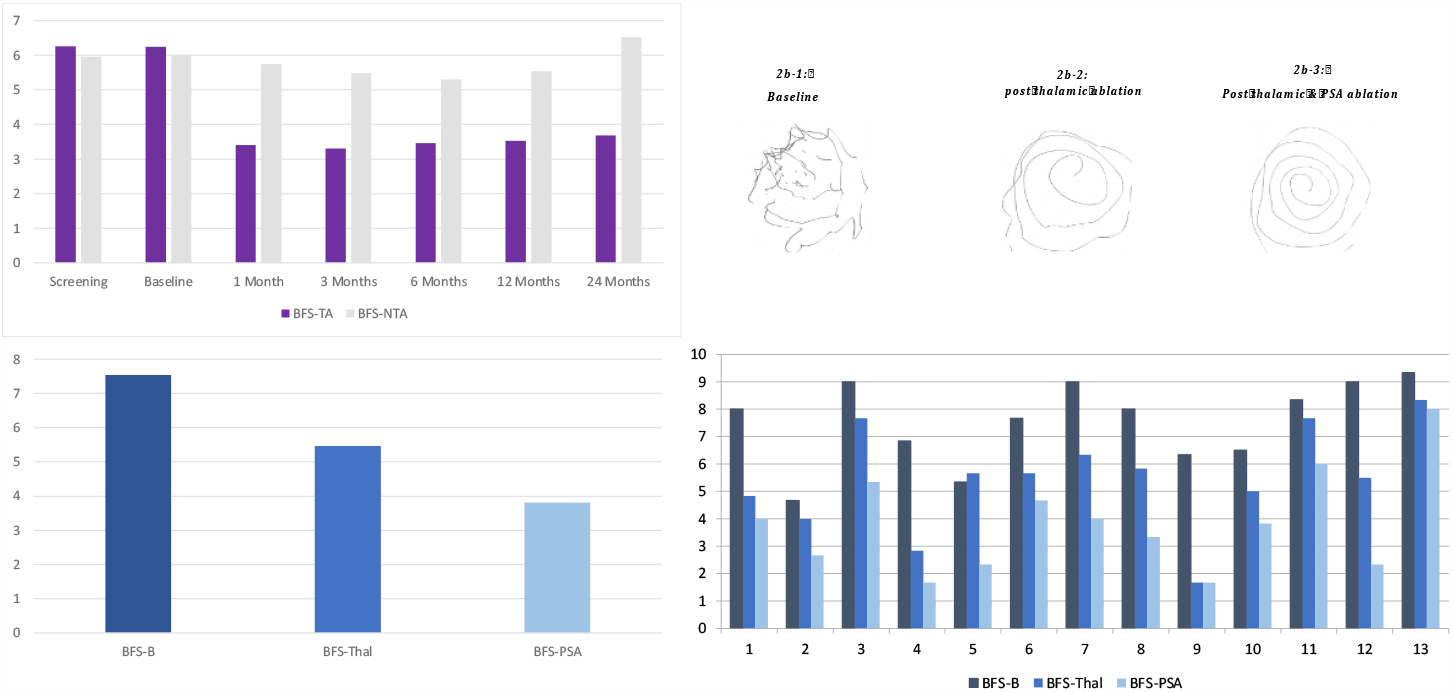
Bain Findley Spiral scores (BFS) ➢ **2a:** The Mean Treated Arm (BFS-TA) and Non-Treated Arm (BFS-NTA) over study period ➢ **2b:** An example of the intra-operative Bain-Findley Spirals (BFS) collected at baseline and after ablation of each target nucleus, demonstrating progressive improvement in tremor: ○ **2b-1:** At baseline, prior to sonications ○ **2b-2:** After Thalamic ablation ○ **2b-3:** After Thalamic + PSA ablation ➢ **2c:** The mean intraoperative BFS scores demonstrating sequential improvement in the mean spiral scores at baseline (BFS-B), after Thalamic (BFS-Thal) and then thalamic + PSA (BFS-PSA) ablations. ➢ **2d:** The mean intraoperative BFS scores for each individual patient, demonstrating the range of improvement from baseline (BFS-B), after Thalamic (BFS-Thal) and then thalamic + PSA (BFS-PSA) ablations.

The mean baseline BFS in the non-treated arm (BFS-NTA) was 6.0 and at 12 months post-MRgFUS was 5.5 (7.7% improvement (Friedman Test: p<0.001) but worsened to 6.5 by 24 months (8.8% worse than baseline (Friedman Test: p<0.001), (Figure 2a).

### Differential effects of Thalamic and then PSA ablation

The blinded assessments of the intra-operative spirals (Figure 2b) at Baseline (BFS-B), after thalamic (anterior-VIM/VOP) ablation (BFS-Thal) and after PSA ablation (BFS-PSA) were compared and showed the following mean percentage improvements (Figure 2c):

- BFS-B to BFS-Thal of 27.9% (Wilcoxon Signed Ranks Test: p < 0.001).
- BFS-THAL to BFS-PSA of 30.1% (Wilcoxon Signed Ranks Test: p < 0.001).
- BFS-B to BFS after Thal and PSA lesions of 49.1% (Wilcoxon Signed Ranks Test: p < 0.001)

These changes were statistically significant across each stage of the procedure (Friedman Test: p<0.001). The individual patient’s BFS-B, BFS-Thal & BFS-PSA scores demonstrate sequential change after lesioning each target nucleus (Figure 2d).

### Secondary Outcomes

The effect of the thalamic and PSA lesions on the secondary outcomes are shown in Table 2. The CRST (including tremor differential scores), QUEST and PHQ scores across the study period are demonstrated in Figures 3 and 4. All measures showed significant improvements (Wilcoxon Signed Ranks Tests: p<0.05), except for the PHQ-9 which showed a non-significant improvement (p=0.145) at 12 months and thus was not collected at 24 months (Figure 4d).

**Table 2:** Secondary outcome measures as mean values at baseline, 12 months, 24 months with percentage change from baseline to time point in brackets (%) and corresponding p value from statistical analysis (Wilcoxon Signed Ranks Test) calculated from baseline to 24month data.

|  | Mean<br>Baseline | Mean<br>12 months | Mean<br>24 months | P value<br>WSRT Baseline<br>to 24 months |
| --- | --- | --- | --- | --- |
| CRST (total) | 58 | 28.8 (-49.5%) | 27.1 (-52.6%) | 0.003 |
| CRST-A Score | 16.5 | 6.5 (-60.7%) | 6.4 (-61%) | 0.003 |
| CRST-B Score | 21.7 | 14.8 (-32.0%) | 15.1 (-30.4%) | 0.004 |
| CRST-C Score | 19.8 | 7.5 (-62.0%) | 6.8 (-65.6%) | 0.002 |
| QUEST (QT) | 87.1 | 54.5 (-37.5%) | 54.2 (-37.8%) | 0.001 |
| PHQ-9 | 6.8 | 4.3 (-36.4%) | Not continued | 0.145 |
| UETTS-TA | 6.8 | 1.79 (-73.9%) | 1.83 (-72.9%) | 0.001 |
| RT-TA | 0.7 | 0.0 (-100%) | 0.2 (-76%) | 0.008 |
| PT-TA | 3.2 | 0.8 (-73%) | 1.0 (-69%) | 0.003 |
| IT-TA | 2.9 | 0.9 (-68%) | 0.7 (-77%) | 0.002 |
| UETTS-NTA | 5.76 | 3.77 (-34.7%) | 4.0 (-30.5%) | 0.003 |

**Figure 3:**
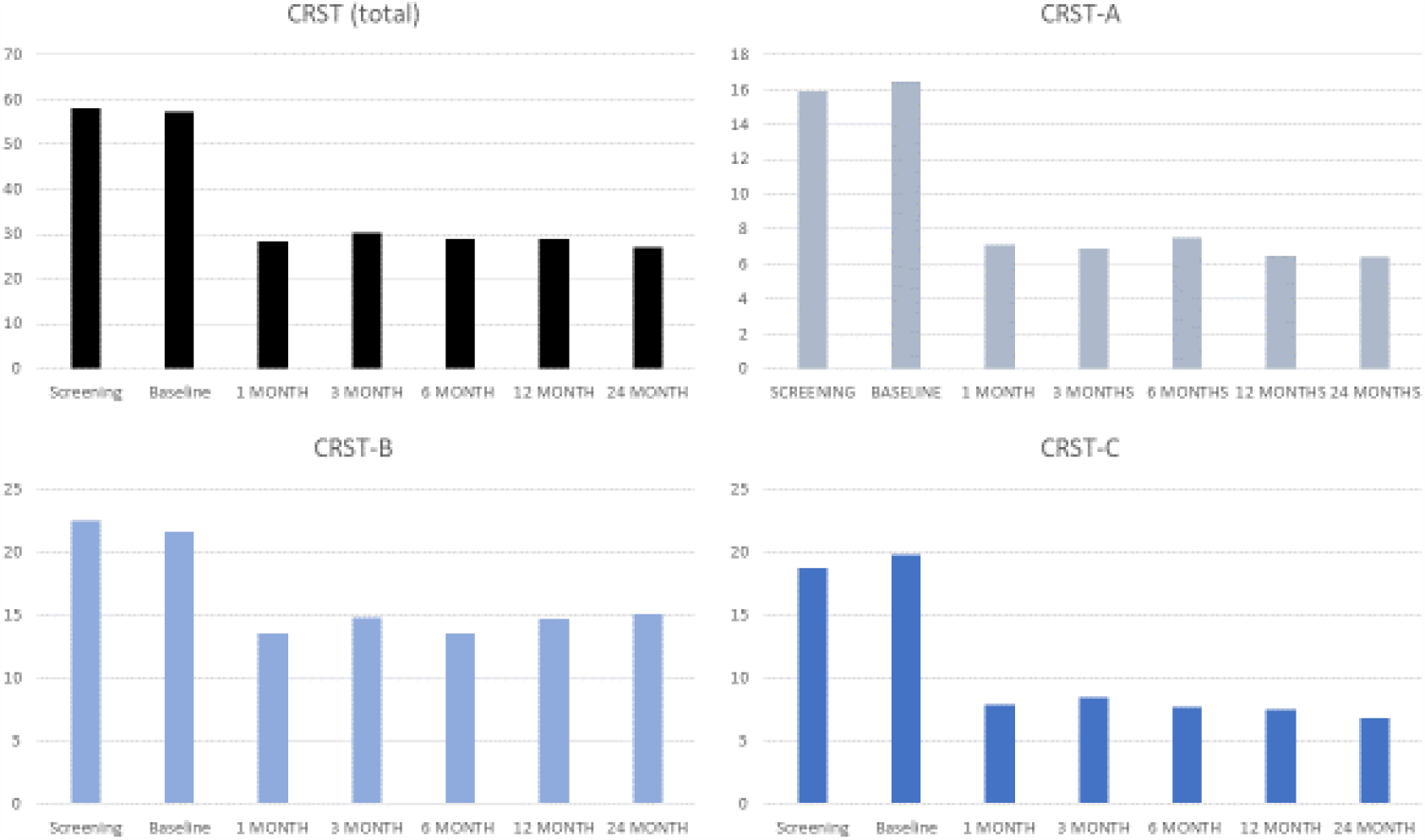
Clinical Rating Scale for Tremor (CRST) ➢ **3a**: Mean CRST total (Parts A+B+C) over the study period. ➢ **3b** Mean CRST Part A: Tremor severity from all affected body parts (bilateral) ➢ **3c**: Mean CRST Part B: Tremor in various manual tasks ➢ **3d**: Mean CRST Part C: Tremor related disability

**Figure 4:**
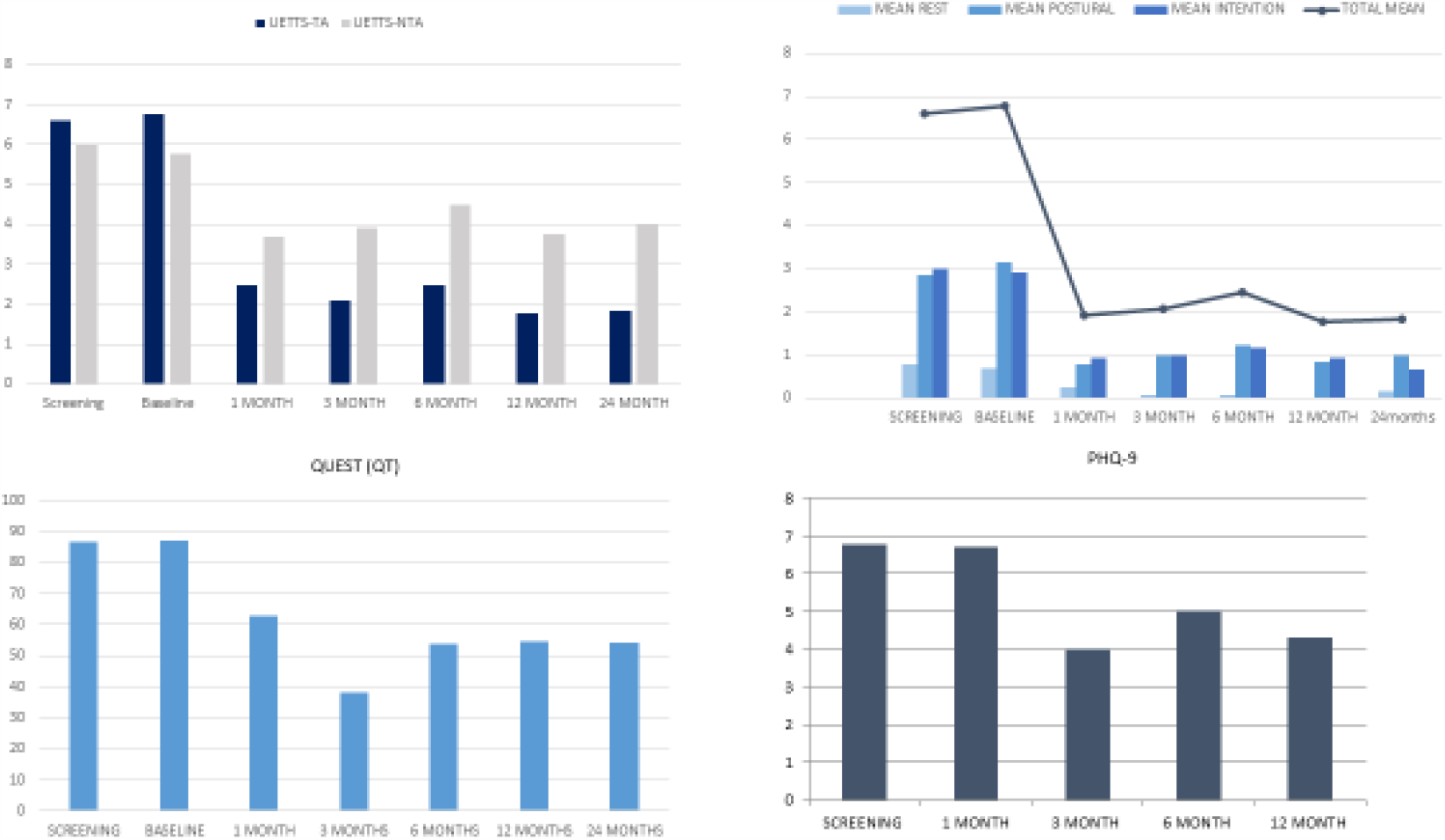
Self-Reported Health Questionnaires and Tremor Differentials. ➢ **4a:** The mean Upper Extremity Total Tremor Score (UETTS) component from CRST-A over the study period, demonstrating both the mean scores for the Treated Arm (UETTS-TA) and the mean score of the Non Treated Arm (UETTS-NTA). ➢ **4b:** Treated Arm Tremor Score over the study period with Total Tremor Score (UETTS-TA) as line graph (black line) and its components Rest Tremor (RT-TA), Postural Tremor (PT-TA) and Intention Tremor (IT-TA) as column graphs (in shades of grey). ➢ **4c:** The mean Quality of life in Essential Tremor questionnaire (QUEST) self-reported Tremor Scores over the study period. ➢ **5d:** The mean Patient Health Questionnaire 9 (PHQ-9) scores, of self-reported depression, over the study period.

The clinican assessed CRST scores improved markedly in all 3 sub-categories

- The mean CRST-A score, a bilateral tremor summation that includes the untreated as well as the treated side (Figure 3b), decreased from 16.5 at baseline to 6.5 at 12 months post MRgFUS), a 60.7% improvement. This was stable at 24 months with the mean CRST-A score 6.4, a 61.0% improvement (Wilcoxon Signed Ranks Test: p=0.003).
- The mean CRST-B score, a bilateral assessment of tremor on specific motor tasks including items from the untreated side (Figure 3c), improved from 21.7 at baseline to 14.8 at 12 months a 32.0% improvement. This was stable at 24 months with the mean CRST-B score 15.1, a 30.4% improvement (Wilcoxon Signed Ranks Test: p=0.004).
- The mean CRST-C score (Figure 3d), an assessment of the impact of tremor on daily function and tremor related disability, was 19.8 at baseline and decreased to 7.5 at 12 months, a 62.0% improvement from baseline to 12 months. This was stable at 24 months with the mean CRST-C score 6.8, a 65.6% improvement (Wilcoxon Signed Ranks Test: p=0.002).

The patient assessed scores also improved markedly

- The Quality of Life in Essential Tremor (QUEST) questionnaire (Figure 4c), mean self-reported tremor score improved from 87.1 at baseline to 54.0 at 12 months, a 37.5% improvement. This was stable at 24 months with the mean QUEST score 54.2, a 37.8% improvement (Wilcoxon Signed Ranks Test: p=0.001).
- The mean Patient Health Questionnaire 9 (PHQ-9) score, a patient self-reported assessment of depression (Figure 4d), improved after MRgFUS from 6.8 at the screening assessment to 4.3 at 12 months, a 36.4% improvement but this change was not statistically significant (Wilcoxon Signed Ranks Test: p=0.145). PHQ-9 was not collated at 24 months follow up.

## Adverse Events

During the procedure: 11 patients experienced pain from the stereotactic frame pins, and 1 developed a pin site haematoma. During sonications a feeling of ‘vertical rotation’ (in which the patients’ legs slowly (over a few seconds) lifted upwards to vertical, pivoting around the neck) (n=7), headache (n=6) and nausea (n=4) were encountered particularly when higher energy was utilized and lasted for the duration of sonication.

The adverse event profile over the course of the study are summarised in Table 3. At 24 months post-MRgFUS, 1 patient had a mildly unsteady gait but walked unaided. Another had persistent mild hemi-chorea but nevertheless felt that he had a net gain from MRgFUS due to effective tremor suppression. This patient’s MRI 12 months after MRgFUS showed a lesion in STN.

**Table 3:** Time frame of post-MRgFUS adverse events encountered in the study.

|  | Treatment<br>Day | Post MRgFUS Treatment (*D= day; M= month*) |  |  |  |  |  |  |
| --- | --- | --- | --- | --- | --- | --- | --- | --- |
|  |  | 1D | 7D | 1M | 3M | 6M | 12M | 24M |
| Fatigue | 3 | 4 | 4 | 4 | 3 | 0 | 0 | 0 |
| Headache | 2 | 0 | 1 | 1 | 0 | 0 | 0 | 0 |
| Migraine | 0 | 0 | 0 | 0 | 0 | 0 | 0 | 0 |
| Paraesthesia | 2 | 0 | 0 | 0 | 0 | 0 | 0 | 0 |
| Dysarthria | 4 | 4 | 5 | 4 | 1 | 0 | 0 | 0 |
| Blurred vision | 0 | 0 | 1 | 0 | 0 | 0 | 0 | 0 |
| Dizziness | 2 | 1 | 2 | 1 | 1 | 0 | 0 | 0 |
| Unsteady gait | 6 | 6 | 5 | 5 | 4 | 2 | 2 | 1 |
| Chorea | 0 | 0 | 2 | 2 | 2 | 2 | 1 | 1 |

## Discussion

### Targeting the Thalamus

In this study, the final thalamic lesions were sited 2.7mm-3.7mm anterior to that commonly performed; many MRgFUS centres deploy A-P co-ordinates that are 25% the length of the AC-PC line anterior to PC. Thus our thalamic lesions straddle the anterior-VIM and VOP border. Our lateral and vertical VIM co-ordinates are similar to those used by other centres. Boutet et al. (2018) demonstrated that, using the 25% of the A-P distance anterior to PC method, their optimal lesions for tremor suppression were situated in the posterior portion of VIM and straddled the border of the ventro-caudalis nucleus (VC).^16^

### Efficacy of anterior-VIM/VOP and PSA lesions

Our results demonstrate that MRgFUS lesions in anterior-VIM/VOP and PSA produced a mean 43.5% improvement in our primary outcome: the change in BFS-TA score from baseline to 12 months post-MRgFUS, which was maintained at 24 months (Figure 2a). The BFS is recommended by the International Movement Disorder Task Force on Tremor and has the advantages of being simple to collect and provides a permanent record of tremor that can subsequently be assessed by blinded assessors.^24^ It is more difficult to diminish tremor present in spirals than the overall tremor scored within the CRST-Part A of the treated arm (UETTS-TA) in which there were mean percentage improvements of 73.9% at 12 months and 72.9% at 24 months (Figure 4a). The mean percentage improvement in the rest and action tremor components in the treated arm were similar 53.6% versus 56.4% (postural tremor decreased by 26.5% and intention tremor components by 31.9%) at 24 months (Table 2 and Figure 4b).

Significant improvements were also found in all 3 subsections of the CRST (Table 2 & Figure 3) and the QUEST tremor score (Table 2 & Figure 4c). The PHQ-9 values were broadly stable, suggesting that anterior-VIM/VOP and PSA MRgFUS lesions do not negatively affect mood (Table 2 & Figure 4d).

The blinded assessments of the intra-operative BFS from the treated arm at Baseline (BFS-B), after thalamic ablation (BFS-THAL) and after PSA ablation (BFS-PSA) showed highly significant improvements across each stage of the procedure, demonstrating that the additional PSA lesion had added benefit for most of our patients (Figure 2c).

The beneficial effect on tremor using our methods are similar at 12 and 24 months post-MRgFUS to those obtained by Halpern et al. (2019) using the (more posterior) conventional VIM approach, as judged from improvements in the ‘hand tremor-motor score’ (HTMS); the primary outcome measure used by Halpern and colleagues.^27^

Comparing our data after the anterior-VIM/VOP lesion only stage with the more common posterior approach (25% of the AC-PC line anterior to PC) suggests that lesions that are more anterior in VIM are less effective than those that are more posterior in VIM; a view which is in accordance with Boutet et al (2018) and likely explains why lesioning PSA is then necessary to achieve comparable tremor suppression to that reported using the standard VIM target by Halpern and colleagues.^16 27^ However, it does raise the question as to whether treating anterior-VIM/VOP is necessary if PSA is then lesioned; an issue worthy of further research.

### Bilateral Effects

Our data revealed an initial bilateral anti-tremor effect from the unilateral anterior-VIM/VOP and PSA lesions. This was evident in the untreated arm (UETTS-NTA) data with 34.7% improvement at 12 months and 30.5% at 24 months compared to baseline (Table 2 & Figure 4a). Although this effect was present in the BFS-NTA scores at 12 months (7.7% improvement) they subsequently worsened by 8.8% compared to baseline at 24 months. It is possible that this was a placebo effect, although this was not seen in the sham arm of a study of MRgFUS for ET.^14^ Alternatively, it is possible that an effect on the mechanisms generating tremor in the ipsilateral arm to the lesion occurred and resulted in transient benefit that was subsequently lost, perhaps by natural progression of the tremor in the non-targetted arm. In this regard unilateral thalamic DBS has been shown to have a long term ipsilateral effect.^28^ Furthermore, animal studies indicate that there is a network of connections between the PSA on both sides and that the PSA has contralateral efferent projections to the intraluminal and higher order nuclei in the thalamus and the upper brainstem in the rat and monkey. ^29, 30^ Extensive connections also exist between PSA and the pedunculopontine nucleus (PPN); the latter having about 50% contralateral descending projections to several key motor centres (brainstem and spinal cord); which might account for a bilateral effect of unilateral THAL/PSA lesions. ^31^

### Adverse Events

The most frequent post-MRgFUS adverse effects were mild dysarthria and an unsteady gait, which typically came on within 24 hours. Dysarthria resolved within 6 months, but mild gait unsteadiness was still present in one patient at 24 months and hemi-chorea, that affected two patients, persisted in one patient to 24 months in a mild form.

The mean movement from final anterior-VIM/VOP to final PSA targets with respect to adverse events are shown in Table 4. Analysis of our data showed a trend towards an increased risk of chorea as targetting is moved more inferiorly and an unsteady gait and dysarthria with more inferior-lateral movements (Table 4). However, as 5 patients who did not develop chorea also had 4-5mm inferior movement from the final anterior-VIM/VOP, the development of chorea may be a *probabilistic* effect caused by oedema and/or lesion encroachment on the subthalamic nucleus (STN) as it did not occur in any of the patients in which the inferior movement from final anterior-VIM/VOP was ≤ 3mm. Hemi-chorea has also been reported in 15% of Parkinson’s patients after STN radiofrequency (RF) lesions.^32^

**Table 4:** The mean movement from the final thalamic to final PSA targets with respect to adverse effects.

| Adverse effect | Mean (range) of movement (mm)<br>from Final VIM to Final ZI targets |  |  |
| --- | --- | --- | --- |
|  | Inferior | A-P | Lateral |
| Chorea | 4.25 (3.5-5) | -0.4 (0-[-0.8]) | 0 (0) |
| Unsteady gait | 3.75 (2.5-5) | 0 (0) | 0.5 (0-1) |
| Dysarthria | 3.58 (2-5) | 0 (0) | 0.2 (0-1) |
| None of the above | 2.88 (1-4) | 0.61 (0-3) | 0 (0) |

*A-P = Anterior- Posterior: negative numbers in this plane indicate anterior movement*

Subsequently, we appreciated that there are technical limitations of MRgFUS when sonicating deeper brain tissue due to inevitable focal spot elongation in the cranio-caudal direction caused by changing the angle of incidence of the sonication beam. This is a separate entity to ‘smearing’ of the sonication spot which typically occurs in the right-left and anterior-posterior directions [P Wragg, MSc. Neurosurgery Applications, InsighTec; personal communication]. Focal spot elongation increases the risk of creating a lesion more inferiorly (in this instance in the STN) the PSA. Since appreciating this issue we adjusted our approach and subsequently treated a further 15 ET patients with MRgFUS VIM/PSA lesions without inducing chorea. Furthermore, in order to diminish oedema, we changed our practice from only administering 4mg of dexamethasone intravenously during MRgFUS to also giving a 5 day oral reducing course afterwards.

The mean lateral movement from the final anterior-VIM/VOP to final PSA target for the patients who developed an unsteady gait or developed dysarthria are shown in Table 4. There are trends for ataxia and dysarthria to occur with more inferio-lateral placements but these adverse effects usually resolved (Table 3).

### Comparison of our results with previous MRgFUS VIM ablation studies

The results of the international multicentre ET002E trial (Halpern et al. 2019) show similar benefits from their VIM only compared to our anterior-VIM/VOP and PSA approach on the ‘hand tremor-motor score’ – the primary outcome measure from the ET002E trial.^27^ Both studies have a mean score of 8 at 12 months and are broadly similar at 24 month follow up (ET002E: 7.5 v 8 in our study but by 24 months 33% of the patients had withdrawn from the ET002E study compared to 7% of our cases).^27^

Comparison of the adverse event profile in our study with that of the ET002E study show that there were fewer persisting adverse effects in our study at 2 years compared to those in the ET002E (15.4% versus 48%) and the profiles differed, as we had no persisting paraesthesia and less ataxia but did have one case of persisting chorea (Table 5a).^27^

**Table 5:** Tables comparing the persisting adverse event profile (AE) in this study with that of published data. ➢ **5a:** Comparison of the persisting adverse event profile (AE) in this study with that of published ET002E study.^27^ The number of persistent adverse effects divided by the number of patients remaining in the relevant study at 2 years is expressed as a percentage.

| Study | Halpern et al 2019 | Jameel et al 2020 |
| --- | --- | --- |
| Total number of patients remaining in study at 2 years post-MRgFUS | 50 | 13 |
| Adverse effect | Number of Patients (% of total study patients) |  |
| Paraesthesia | 11 (22%) | 0 (0%) |
| Unsteady gait* | 7 (14%) | 1 (7.7%) |
| Slow movements | 1 (2%) | 0 (0%) |
| Dysmetria | 1 (2%) | 0 (0%) |
| Muscle weakness | 2 (4%) | 0 (0%) |
| Taste Disturbance | 1 (2%) | 0 (0%) |
| Fatigue | 1 (2%) | 0 (0%) |
| Chorea | 0 (0%) | 1 (7.7%) |
| Total no. of AEs at 2 years | 24 (48%) | 2 (15.4%) |

*\* For simplicity ataxia, unsteadiness and imbalance have been grouped under 'unsteady gait'.*

**➢ 5b:**
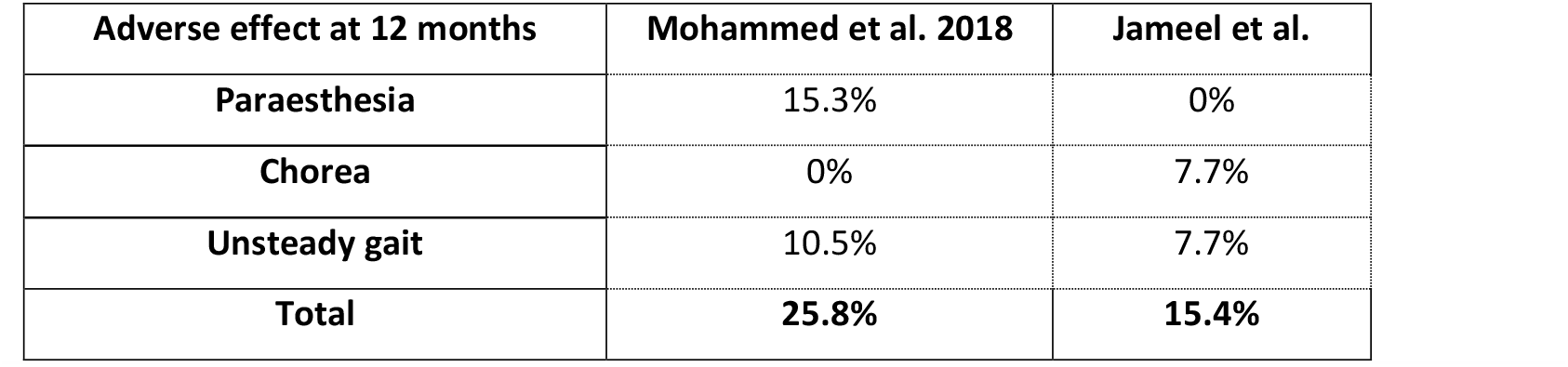
Comparison of the persistent adverse effects at 12 months post-MRgFUS in our study with that of a meta-analysis of MRgFUS treatment of essential tremor.^17^

| Adverse effect at 12 months | Mohammed et al. 2018 | Jameel et al. |
| --- | --- | --- |
| Paraesthesia | 15.3% | 0% |
| Chorea | 0% | 7.7% |
| Unsteady gait | 10.5% | 7.7% |
| Total | 25.8% | 15.4% |

Furthermore, comparison of the persistent adverse effects in our study with those reported at 12 months in a meta-analysis of MRgFUS for ET, shows a favourable sensory adverse effect profile in our study, most likely attributable to the previously described more anterior VIM/VOP targeting (Table 5b).^17^ In the metanalysis paraesthesia (15.3%) and unsteady gait (10.5%) were the main issues, whereas at 12 months we had 0% paraesthesia and 7.7% unsteady gait. Of note more posterior VIM lesions are associated with a 38 times higher likelihood of adverse sensory effects if they encroach on VC.^16^

### Deep Brain Stimulation

MRgFUS is not associated with the 1.2% risk of symptomatic intracranial haemorrhage attached to DBS nor the multiple ongoing health risks associated with an implanted DBS systems.^33, 34^ Consequently, it is reasonable to infer that MRgFUS is safer than DBS. DBS does the advantage of longer experience in both unilateral and bilateral treatments for tremor. ^41^ Current MRgFUS practice is for unilateral procedures to treat tremor in one arm. There has been some hesitancy to perform bilateral MRgFUS treatments due to historic adverse events associated with Radiofrequency Ablations (RF) for ET, including a significant incidence of dysarthria in patients undergoing unilateral (4.5%) and bilateral (13.9%) RF ablation and to a lesser degree DBS (even after reprogramming) for ET. ^34, 35, 41^ However, recent pilot studies have reported that it is safe to do staged bilateral MRgFUS lesions and thus staged bilateral treatments are likely to become more widely adopted. ^42, 43^

The long-term stability of the anti-ET effect of MRgFUS is unknown, as the only published 5 year data has just 2 patients remaining in the study, whilst DBS has been deployed for several decades.^36^ However, tolerance to stimulation and/or natural worsening of tremor develops, so that the benefit of DBS diminishes by 5 years.^37 38^ It remains to be seen whether MRgFUS will be a more appropriate treatment than DBS for elderly people with ET. The commercial cost of performing MRgFUS at our institution is 51% of that for DBS and the subsequent cost of care is considerably less.

### Limitations of our study

Our study has several weaknesses. First, although the BFS was scored by blinded assessors there was no randomization to a control arm or sham MRgFUS. Secondly, our study is small although the beneficial results on tremor are nevertheless highly statistically significant. Thirdly, it would have been preferable to target anterior-VIM/VOP and the PSA from DTI MR scans, which were not available to us at the time we performed the MRgFUS procedures.

## Conclusion

This study confirms the effectiveness of unilateral anterior-VIM/VOP and PSA MRgFUS for alleviating ET, with a profound contralateral and small transient ipsilateral effect. We show that placing lesions in a more anterior-VIM/VOP and PSA provides similar tremor suppression to that attained by lesions in the conventional posterior VIM target and resulted in a lower incidence of persistent adverse effects and in particular a complete absence of parasthesiae.

## Data Availability

The data up to 2 years post MRgFUS is available on request.

## Acknowledgement

We would like to thank Paul Wragg, Michela Mazzola, Tina Stoycheva and Lesley Honeyfield for their assistance with this study.

## Appendices

### Appendix 1: Inclusion and Exclusion Criteria

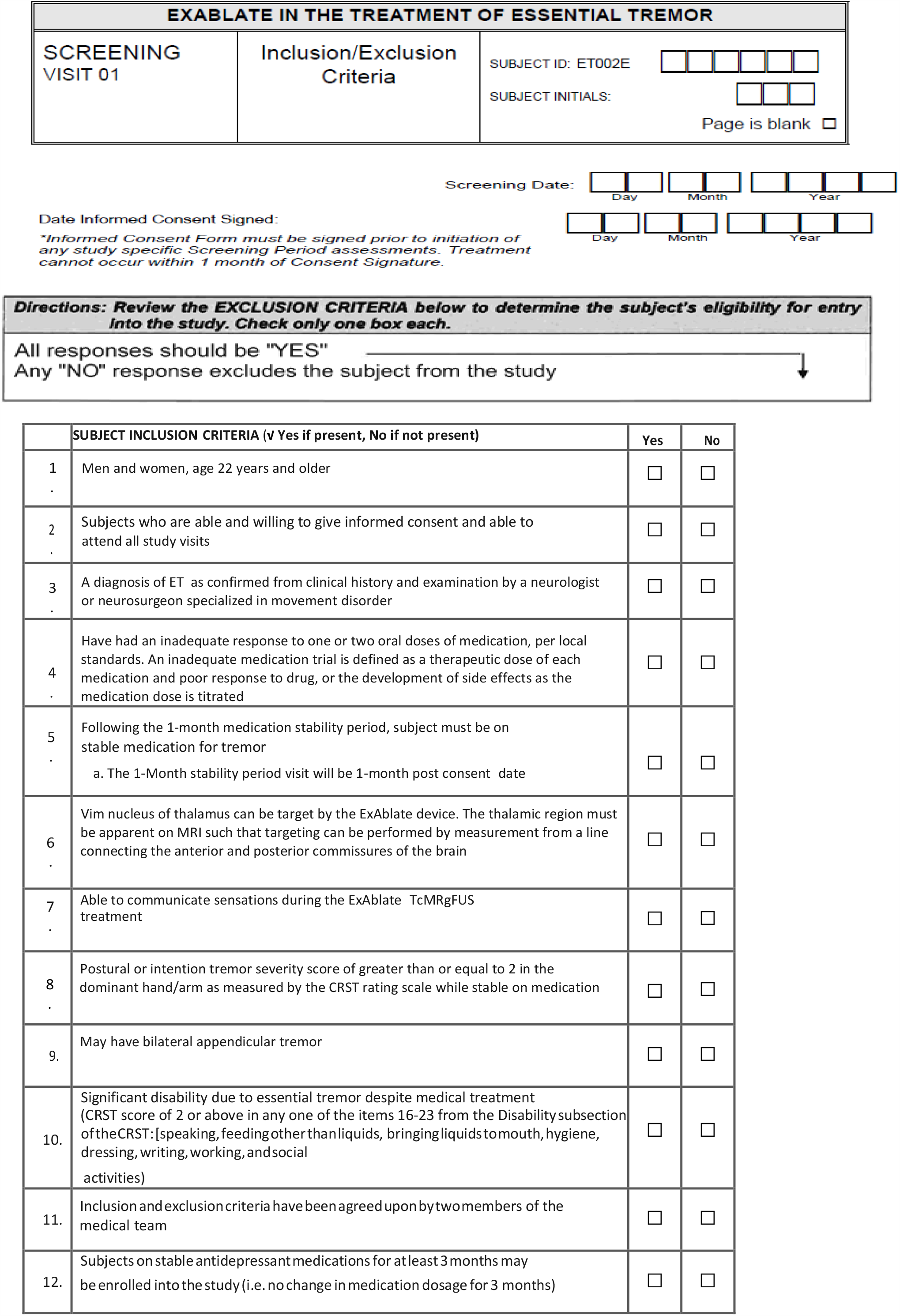

### Appendix 2: Clinical Rating Scale for Tremor (CRST)

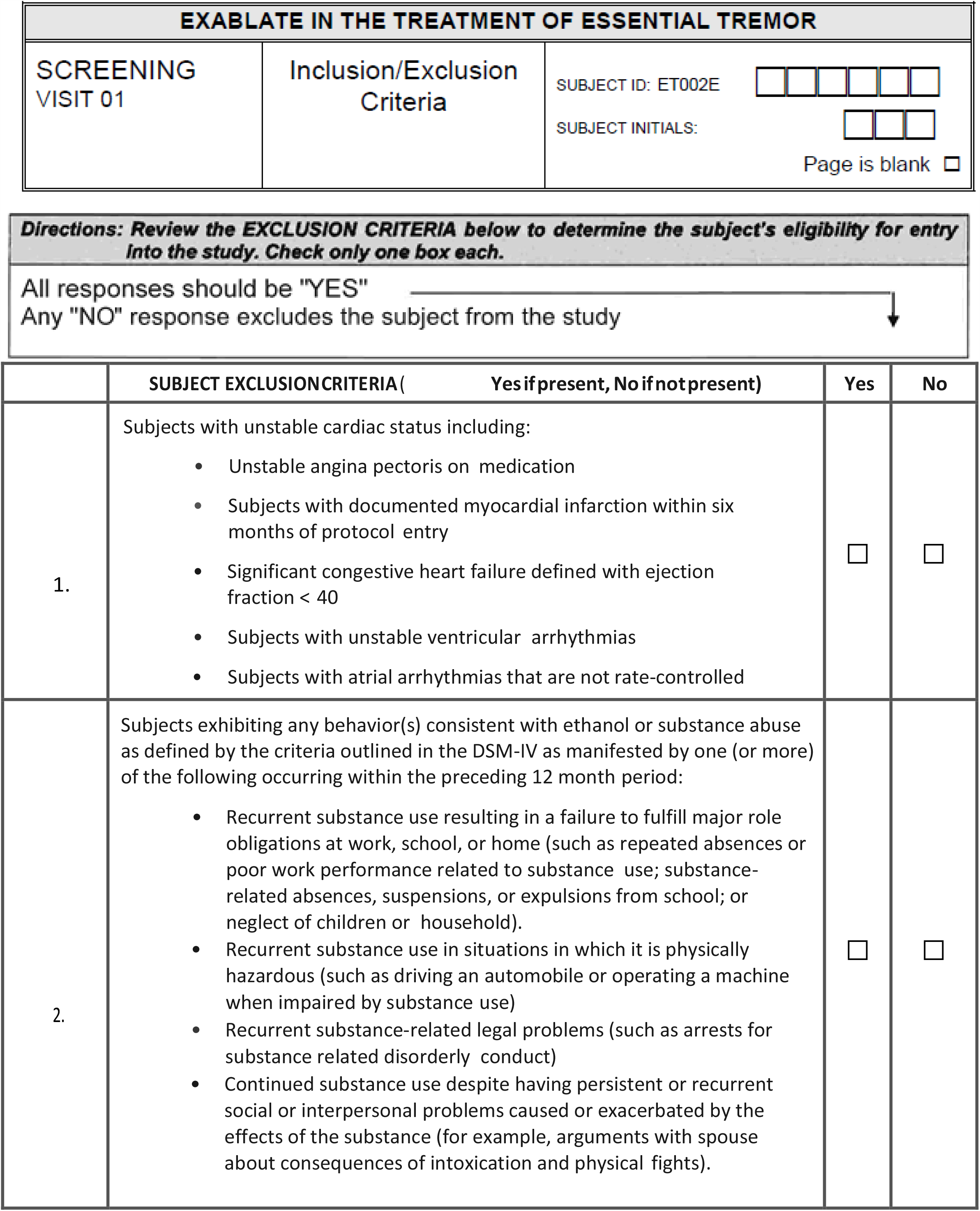
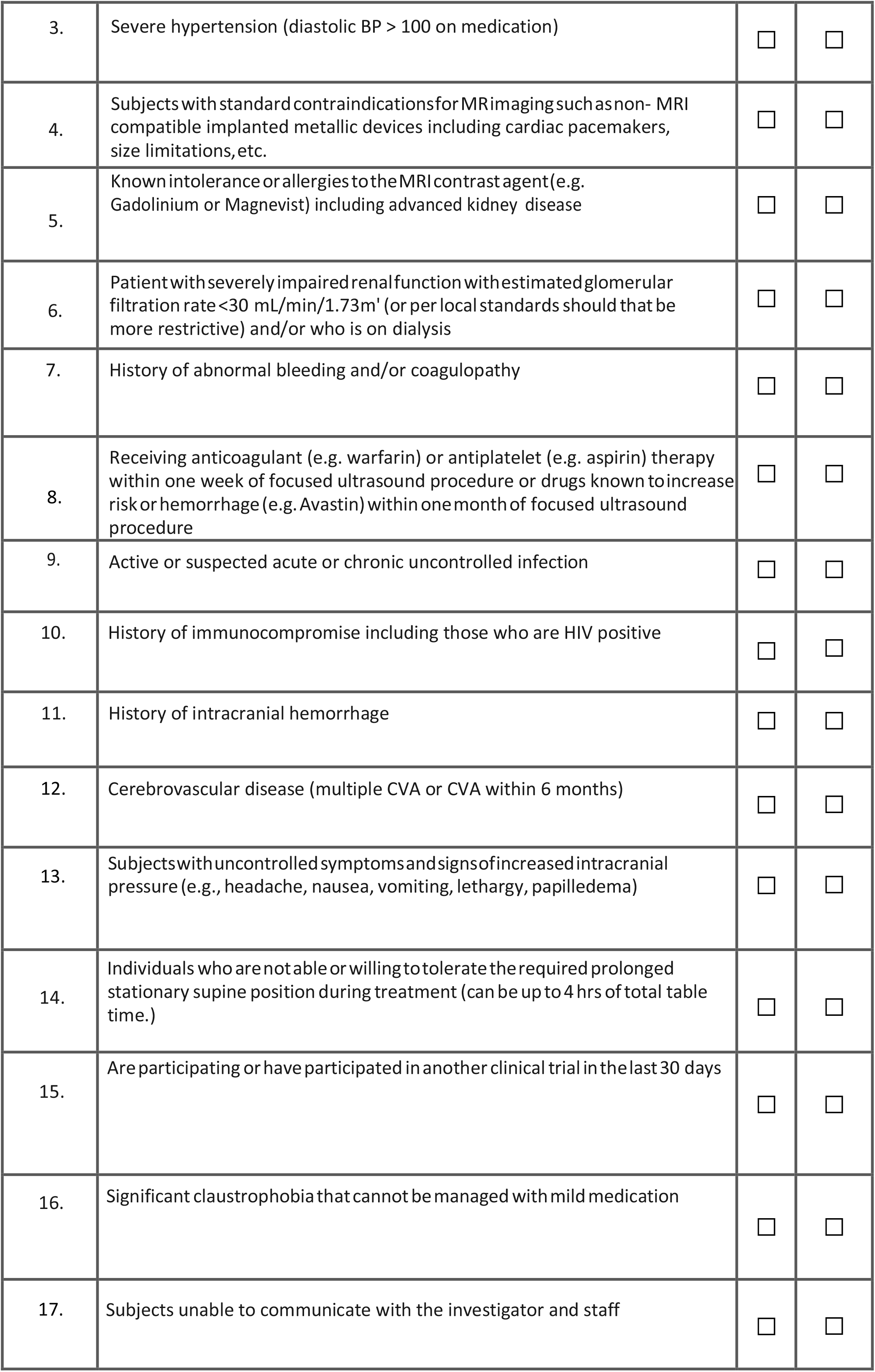
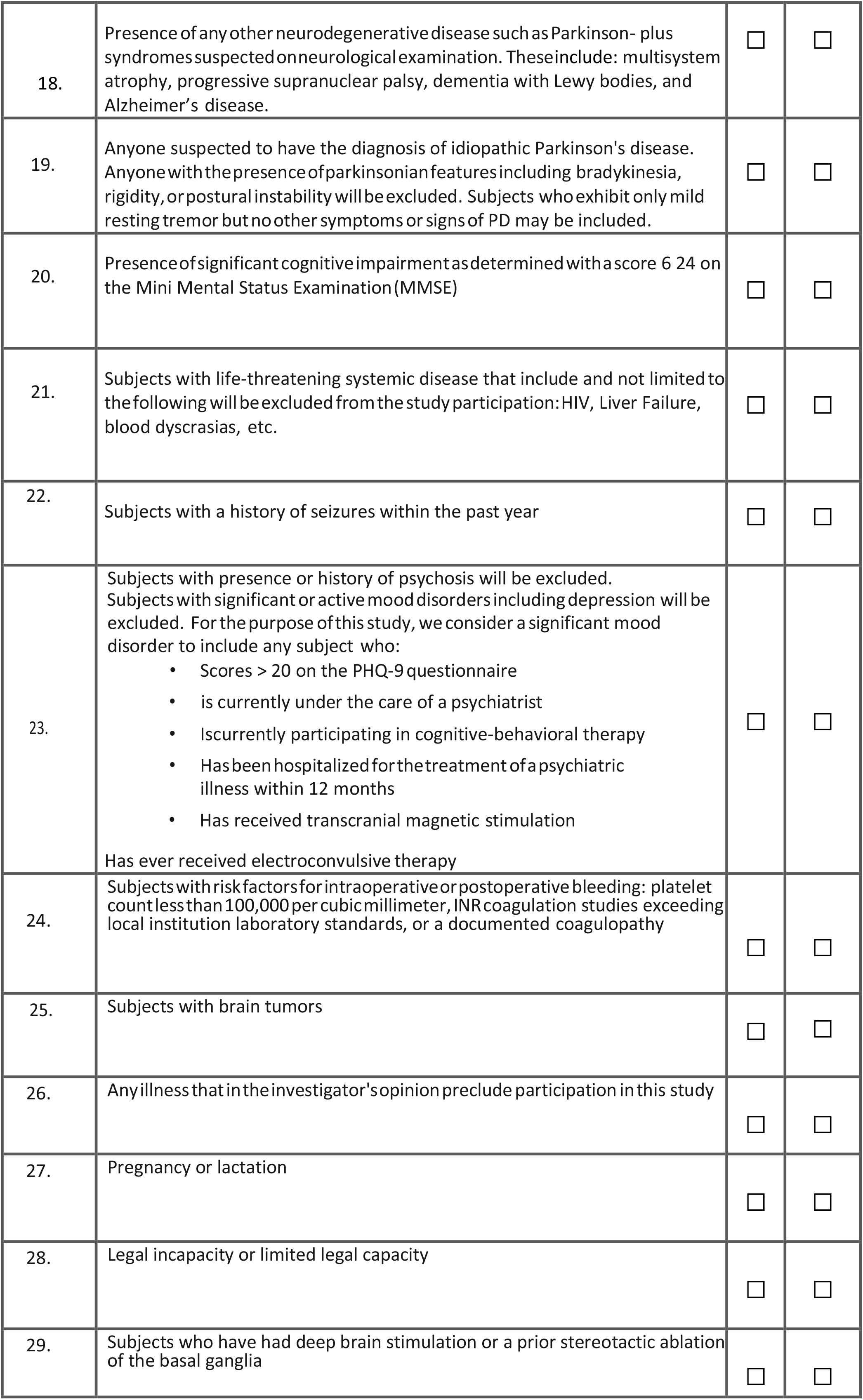
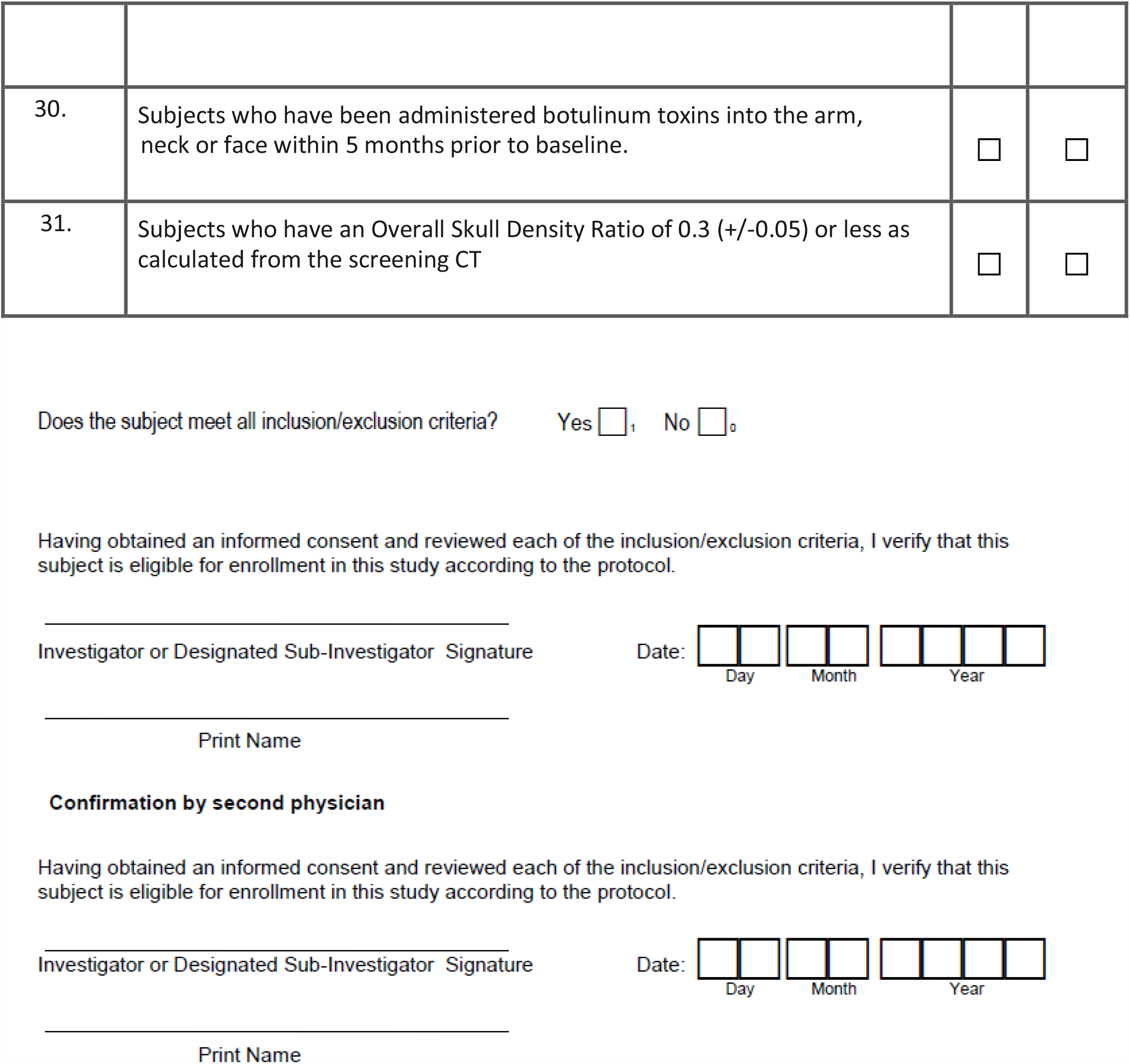
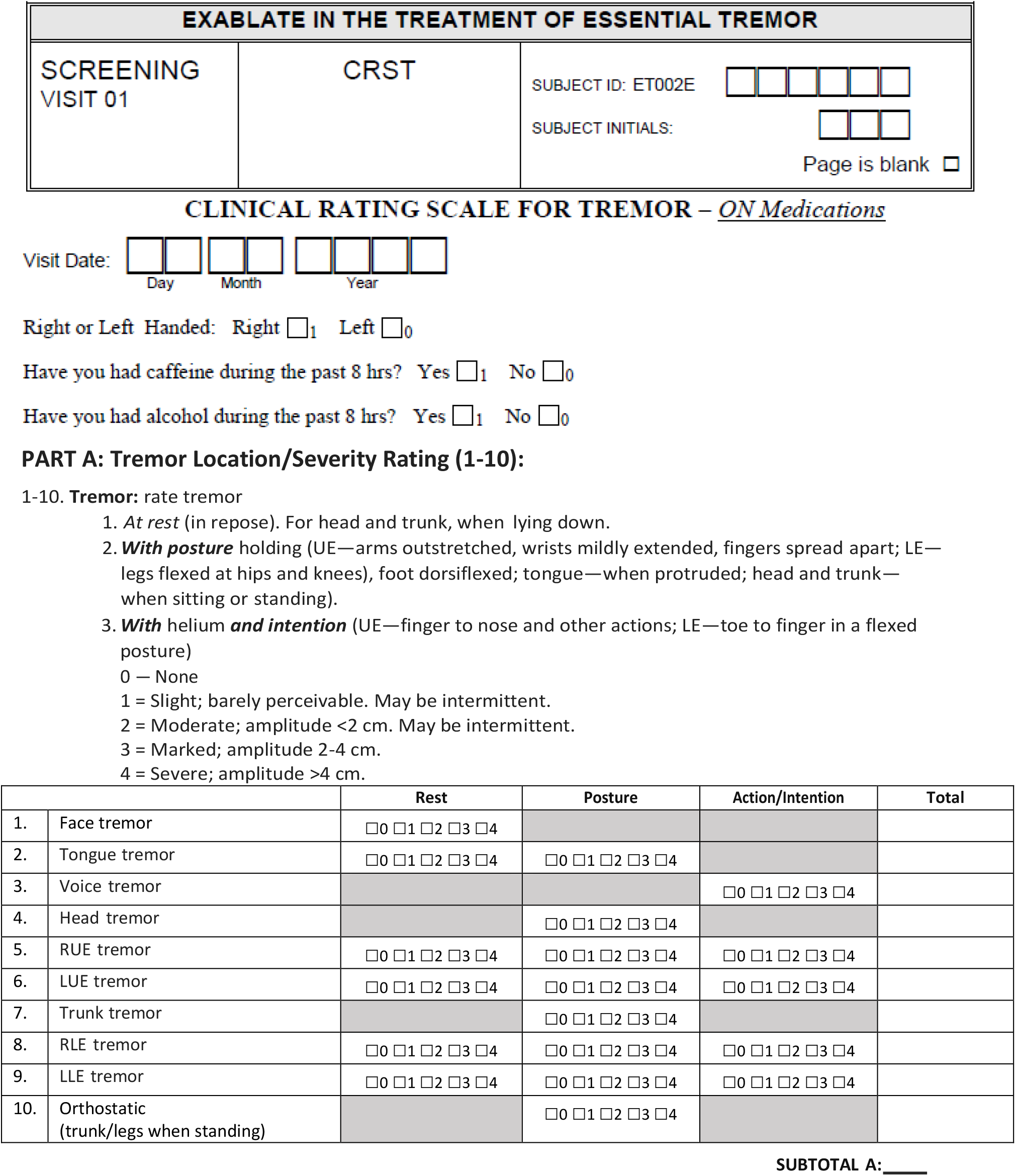
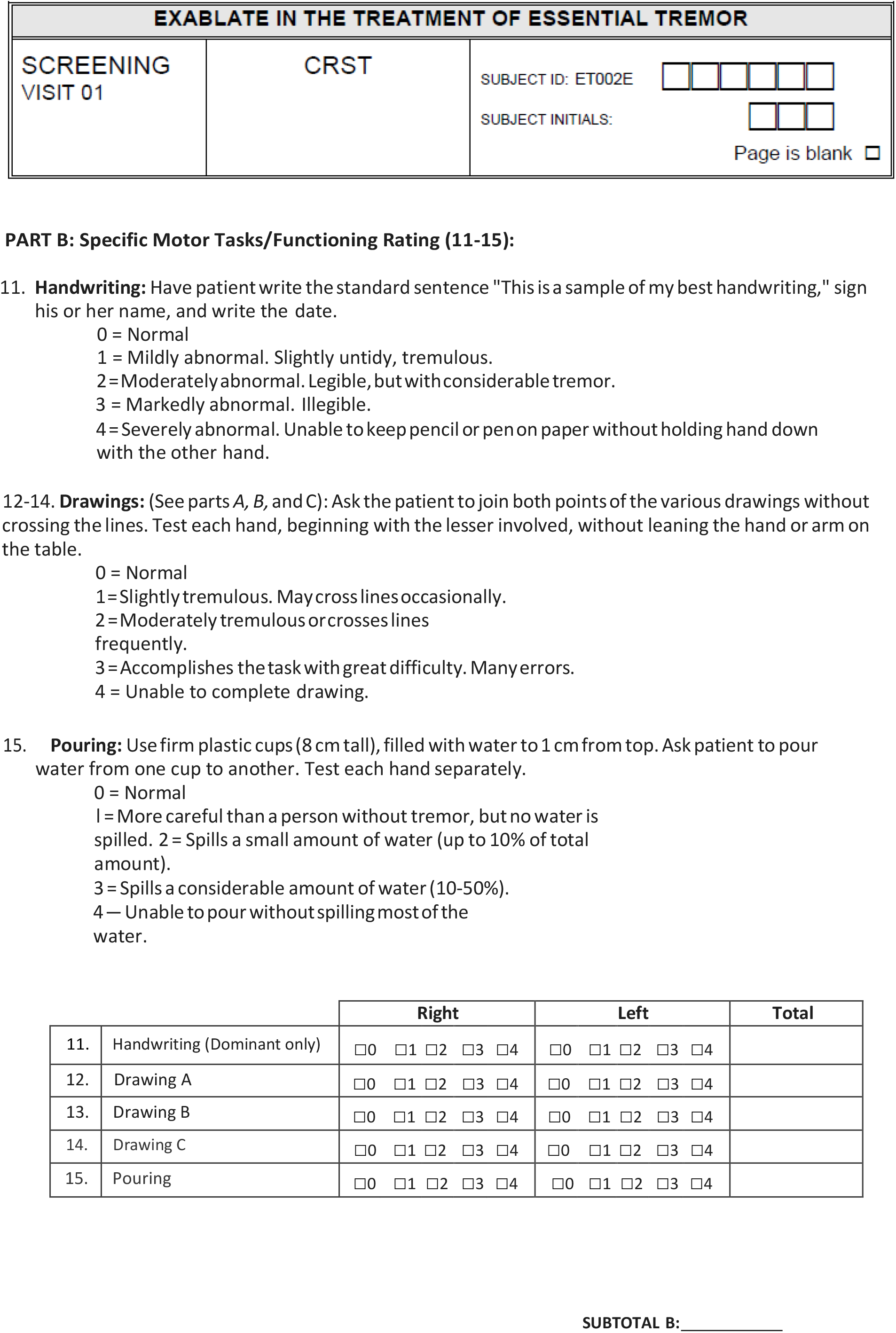
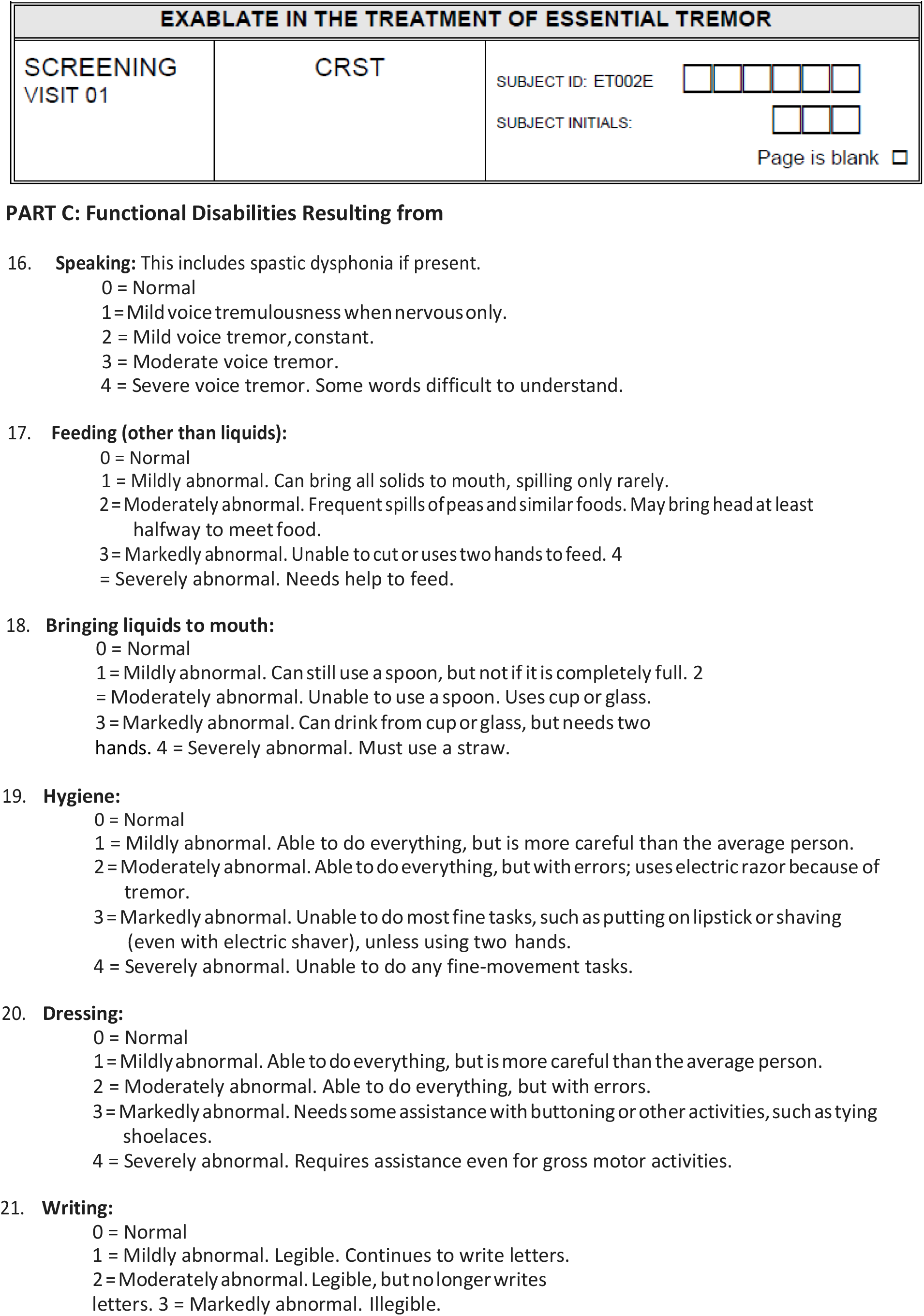
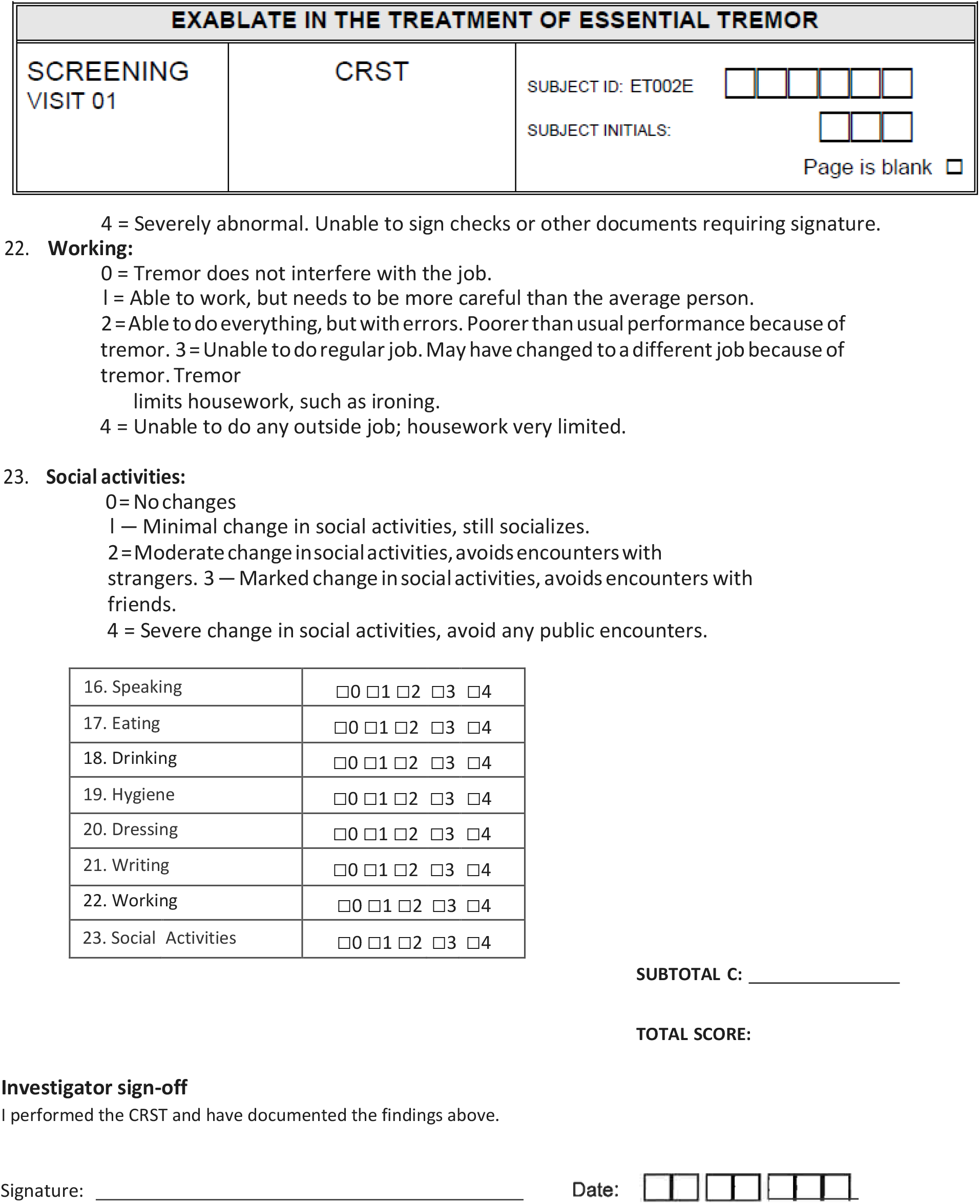
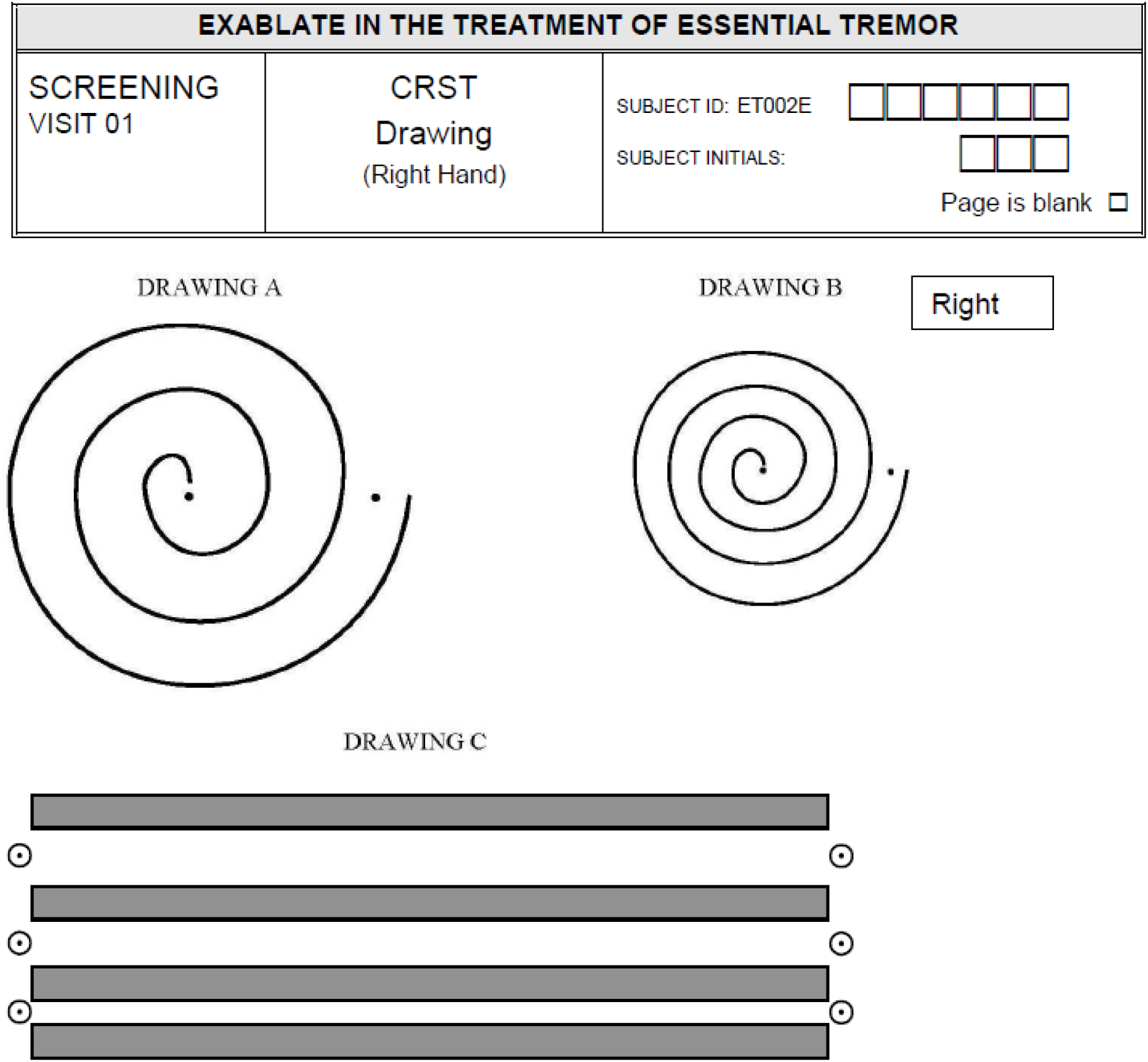
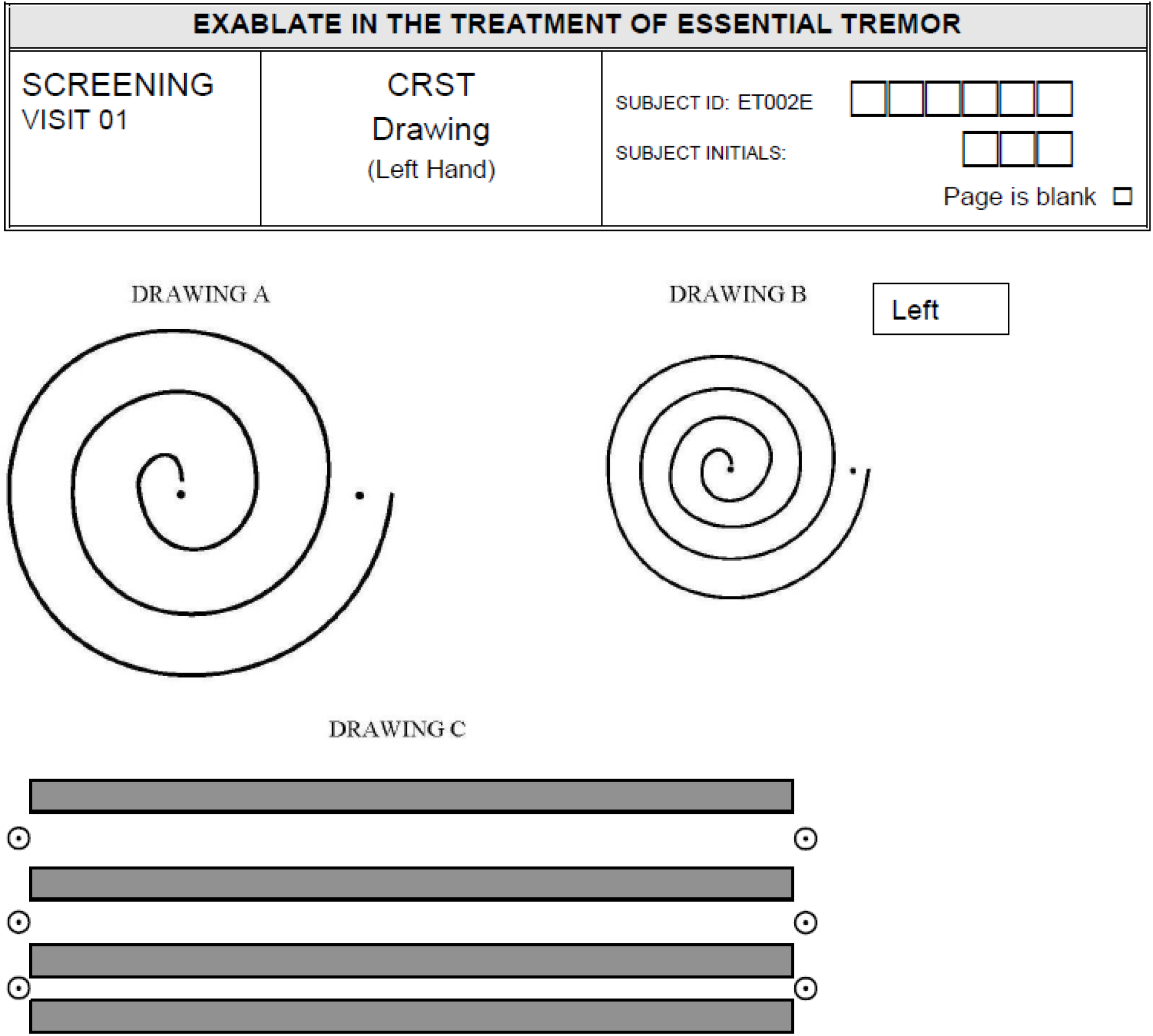
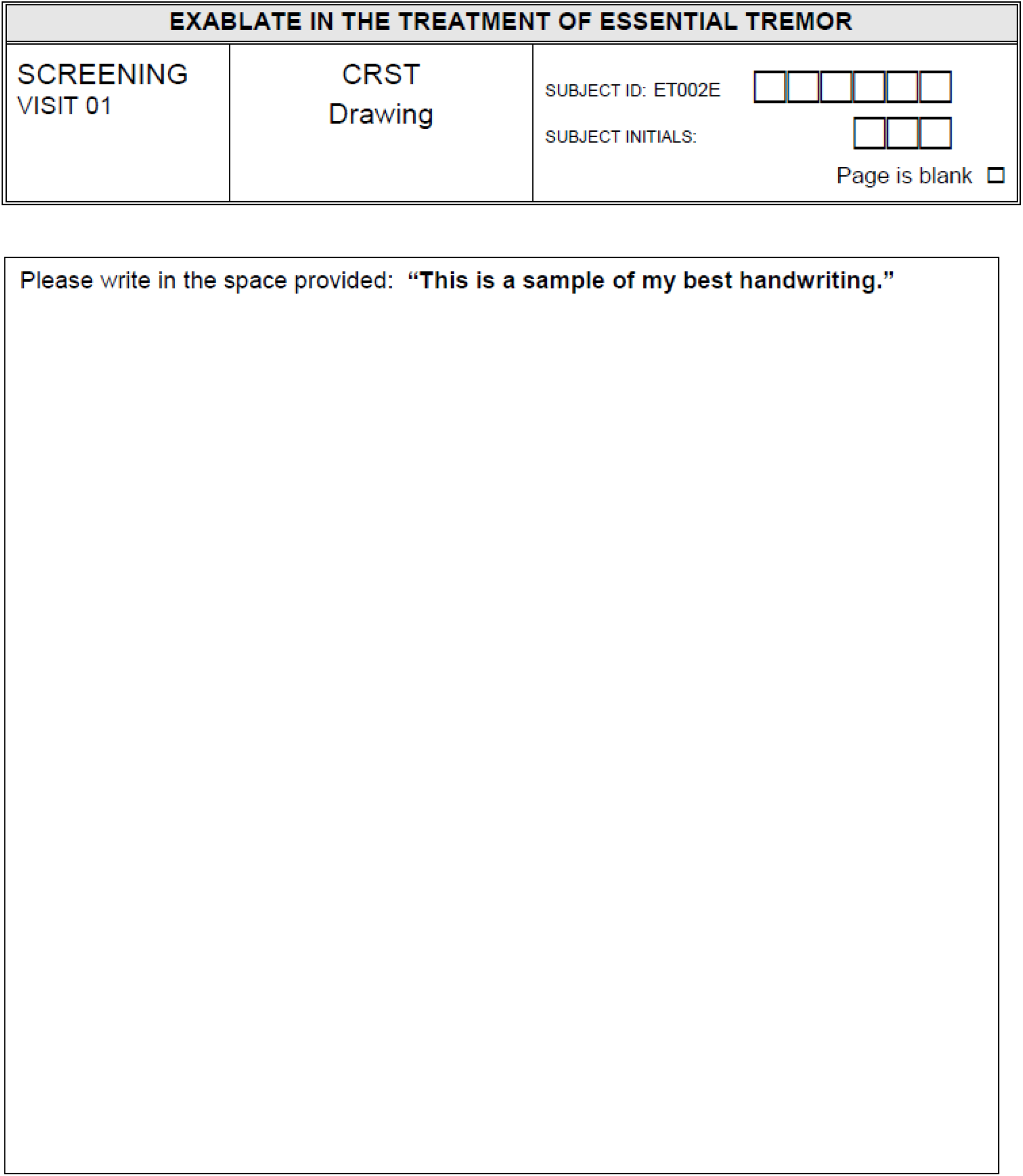

### Appendix 3: MRgFUS treatment

The skull density ratio (SDR), which is the ratio of cortical to cancellous bone, was calculated using a planning CT scan^11^. A low SDR indicates a skull vault that is more absorbent of focused ultrasound, making it more difficult to achieve therapeutic temperature levels at the intracerebral target site; preventing an effective lesion. The lower SDR ‘cut-off’ for exclusion in this trial was < 0.3 (+/- 0.05).

On treatment day, patients were re-assessed to ensure there were no contraindications to treatment with particular care to check that anticoagulant medications had been stopped appropriately. Written informed consent was obtained prior to the procedure. The patient’s head was examined and shaved before fixation of the stereotactic frame (Integra) with local anaesthetic as required. A flexible silicone membrane was secured around the patient’s head and sealed to allow chilled degassed water to circulate and maintain scalp cooling.

MRgFUS was performed using the ExAblate Neuro 4000 device (InsighTec plc Haifa Israel) with a 1024 element phased array transducer operating at 650 kHz. Precision imaging and thermal mapping was performed using a 3T MR (General Electric Milwaukee USA). The procedure took between 4 and 6 hours to perform and involved three stages:

#### Identifying the Thalamic and PSA targets

Using the preoperative MRI study, stereotactic planning of the target was performed. The anterior VIM/VOP was initially targeted at 3mm posterior and 13-15mm lateral to the mid-point between the anterior commissure (AC) and posterior commissure (PC) along the AC-PC line (Figure 1a). This target was then cross correlated with the distance from the third ventricle and the internal capsule. The optimal distance from the wall of the third ventricle is 11-11.5mm and from the lateral border of the thalamus is 2mm. The latter is based on the known motor somatotopy of the ventrolateral thalamic nuclei wherein the homunculus is mapped medial to lateral in the order of face, arm and then leg.

Trial and treatment sonications (see below) were performed to ensure tremor suppression was achieved by tailoring targeting to individual neuroanatomy and sonication parameters to individual neuromodulatory response. Once a satisfactory lesion was placed in the thalamus (thus achieving good control of distal hand tremor) then, providing a clinically significant tremor remained in the treated hand, the target was switched to the PSA. The rationale for using the PSA is supported by published literature from DBS treatment of ET.^40^ The location of the PSA varies slightly between patients (as is true of all the common functional stereotactic targets in clinical practice). Our standard approach to PSA targeting was movement 3 mm inferior, 0.5mm posterior and 0.0mm lateral to the anterior-VIM/VOP target. (Figure 1a).

#### Trial sonications

Low power sonications were initially performed to induce reversible non-permanent lesions using sub-ablative temperatures up to 44°C at the target and confirm the accuracy of the focus location with combined MR imaging and MR thermographic imaging. After each sonication, a brief clinical and tremor assessment including a spiral drawing was performed by a neurologist (PB).

Intravenous paracetamol and ondansetron were administered routinely and fentanyl given intravenously when necessary for more severe pain. The patient’s heart rate, blood pressure, respiratory rate and oxygen saturations were continuously monitored throughout the procedure.

#### Treatment sonications

Once target location and safety were confirmed by trial sonications, further sonications of gradually increasing power were applied to the target in a stepwise manner to ensure permanent ablative temperatures were reached. Ablation occurs when the maximum temperature reaches ≥ 55° Celsius for 1 second, measured by MR thermography. Treatment sonications last between 13-24 seconds and typically are in the range of 10,000-30,000 Joules.

Further sonications were performed after each of which there was a brief interlude that allowed the scalp to cool, and the patient to be reassessed. Treatment sonications were then continued until optimal control of the tremor was achieved. Accurate targeting and successful thermal ablation at the tremor specific nuclei produced immediate tremor reduction, which was visible to both patient and clinician. Sequential spirals drawn whilst in the scanner readily demonstrated progressive tremor reduction.

After the final sonication and subsequent clinical assessment, the stereotactic frame was removed, and the patient re-examined for adverse effects. An immediate post procedure MR brain scan was then performed. All the patients were observed overnight and discharged the day after the procedure and then followed up regularly according to the study protocol.

### Appendix 4: Quality of Life in Essential Tremor (QUEST)

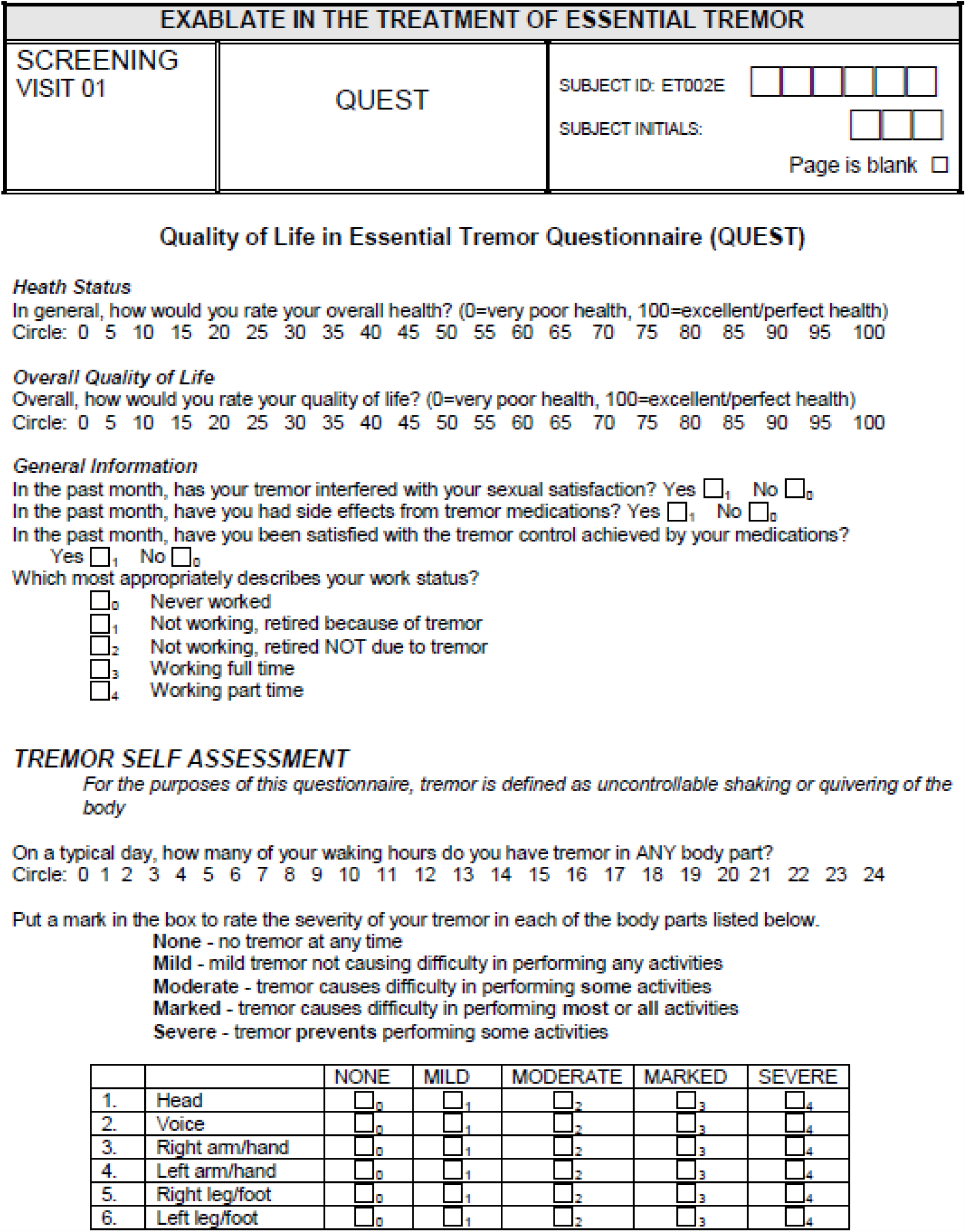
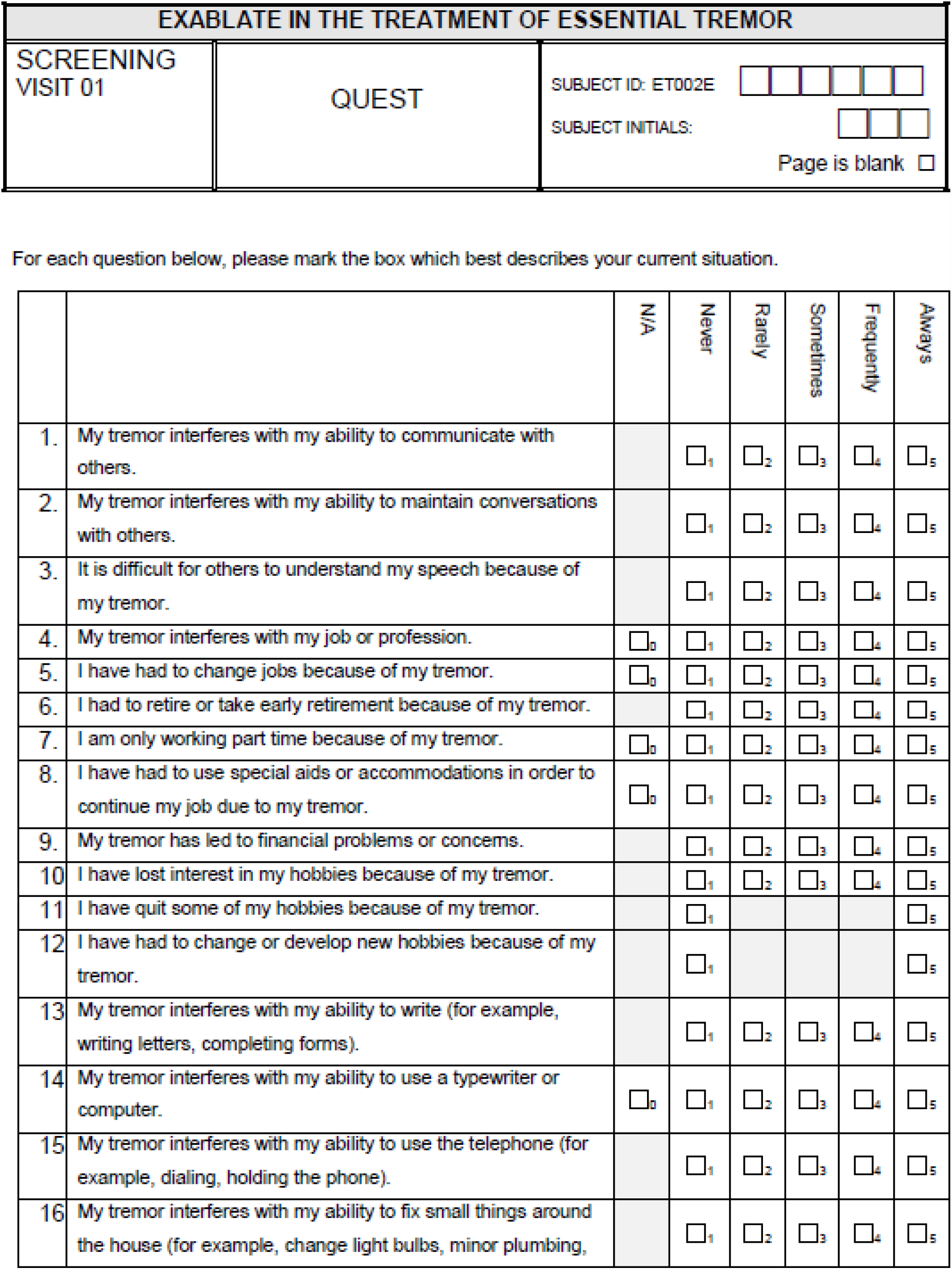
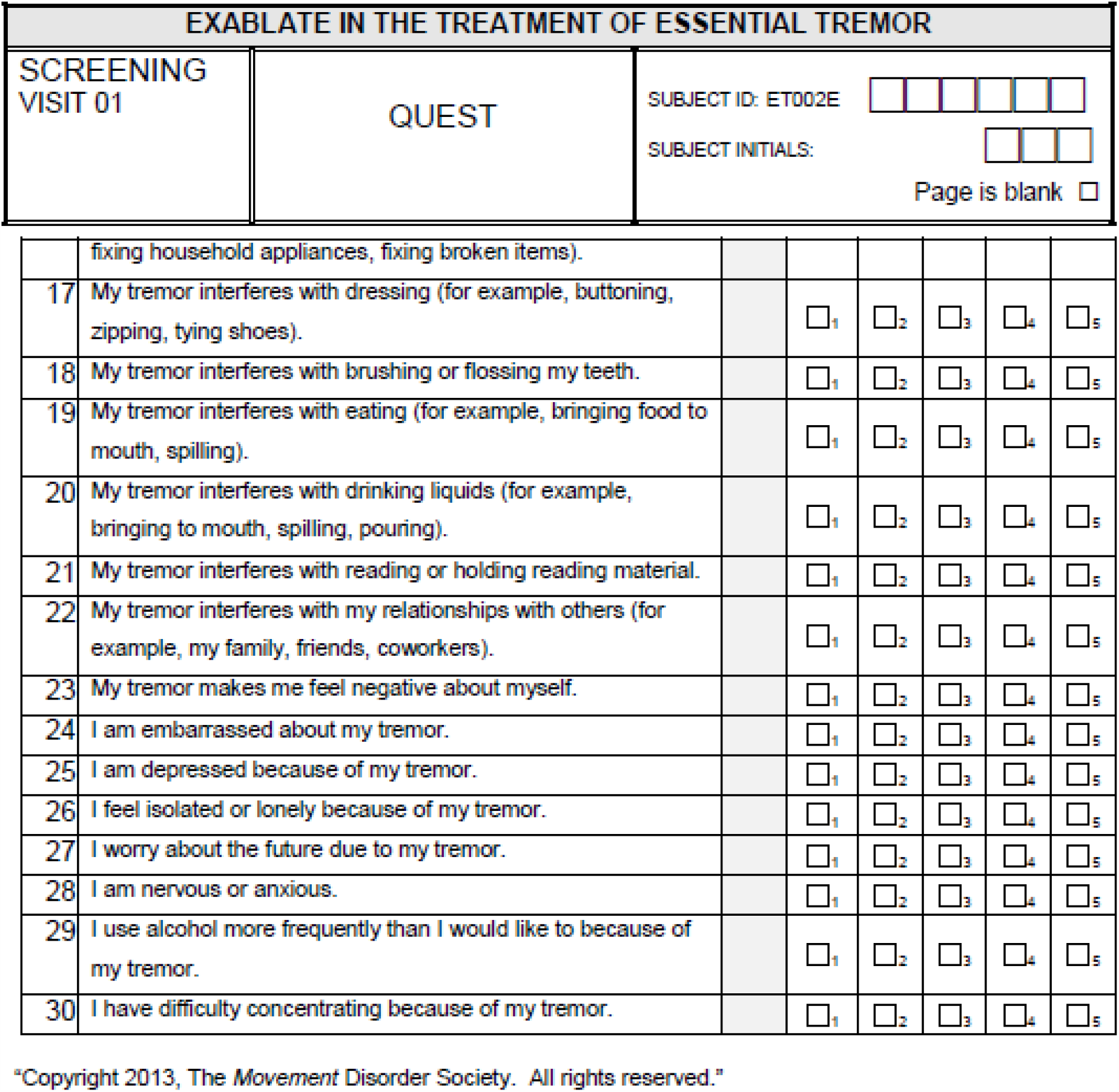

### Appendix 5: Patient Health Questionnaire Nine (PHQ-9)

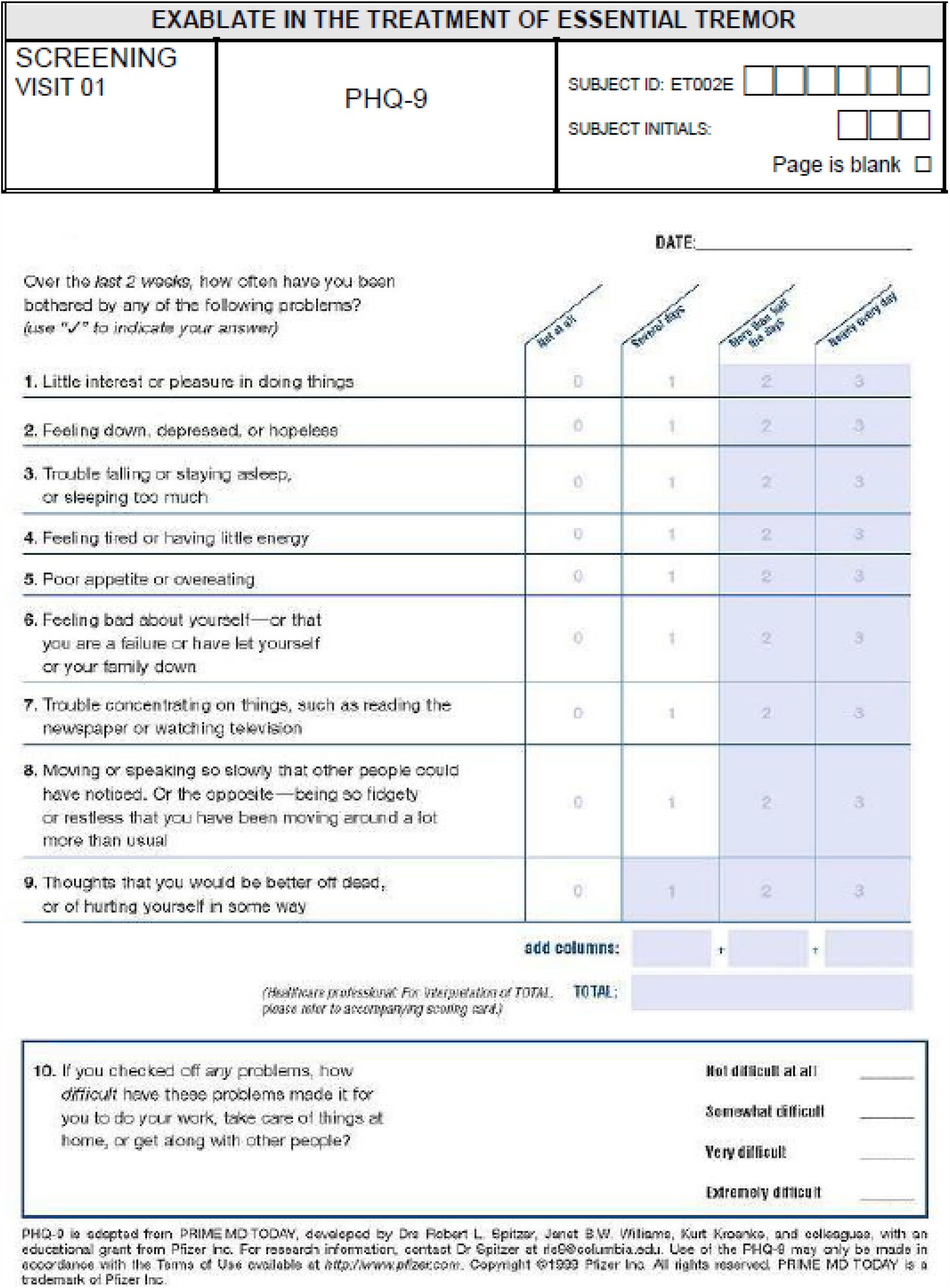

### Appendix 6: Flow chart demonstrating patient numbers at recruitment and completion of study

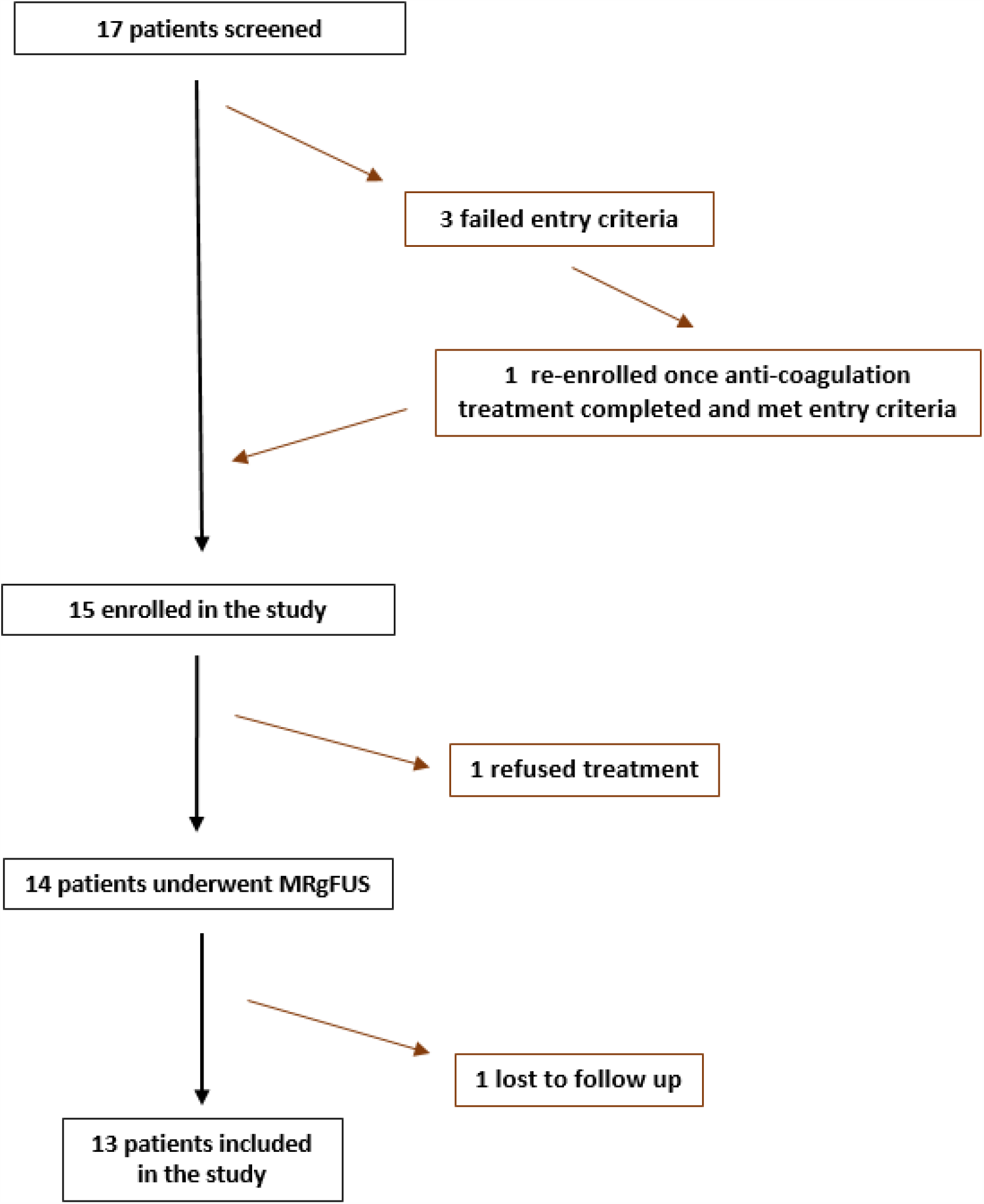

